# TILseg: Automated Whole Slide-Level Spatial Scoring of Tumor-Infiltrating Lymphocytes Reveals Prognostic Patterns in Triple Negative Breast Cancer

**DOI:** 10.64898/2026.01.08.26343727

**Authors:** Lisa L. Carr, Abishek Sankaranarayanan, Khanh Ha, Meenal Rawlani, Anum S. Kazerouni, Jennifer Specht, Laura C. Kennedy, Daniel Reiter, Suzanne Dintzis, Daniel S. Hippe, Mark R. Kilgore, Lynn Symonds, Savannah C. Partridge, Shachi Mittal

**Affiliations:** Department of Chemical Engineering, University of Washington, Seattle, WA 98195, USA; Department of Laboratory Medicine and Pathology, University of Washington School of Medicine, Seattle, WA 98195, USA; Department of Radiology, University of Washington, Seattle, WA 98195, USA; Division of Hematology and Oncology, University of Washington School of Medicine, Seattle, WA 98195, USA; Clinical Research Division, Fred Hutchinson Cancer Center, Seattle, WA 98109, USA; Department of Medicine, Medical Center, Vanderbilt University, Nashville, TN 37232, USA; Department of Bioengineering, University of Washington, Seattle, WA 98195, USA

## Abstract

Stromal tumor-infiltrating lymphocytes (sTILs) are promising biomarkers for predicting therapeutic outcomes in triple-negative breast cancer (TNBC), with higher sTIL levels correlating with improved chemotherapy response and survival outcomes. Currently, sTILs are manually evaluated by pathologists, which is prone to inter-reader variability. In this study, we have developed an AI-driven TIL segmentation pipeline to process entire diagnostic hematoxylin-and-eosin-stained whole slide images for reproducible scoring (global TILseg scoring) and reliable prognostication. This pipeline was optimized and tested using two independent TNBC patient cohorts (n = 57 in the discovery cohort, n = 43 in the validation cohort) with clinical outcomes and follow-up data. The global scores generated by TILseg showed moderate to high concordance with expert scoring (Spearman R = 0.84-0.89) and improved patient stratification (p-value = 0.0191) as compared to manual scoring (p-value = 0.0663). Additionally, we investigate how the spatial localization of sTILs (spatial TILseg) impact survival outcomes by identifying TILs in selected stromal subsets (0.02-2 mm from the epithelial clusters). Our findings have shown that TILs up to 50 µm from epithelial regions prove to be most prognostic in predicting recurrence-free survival post-neoadjuvant chemotherapy with higher statistical significance than both manual and global TILseg scoring. Further, spatial TILseg scoring was more significantly associated with pathological complete response status in both patient cohorts. In summary, we present an AI-based digital tool for robust sTIL scoring and spatial mapping to enhance its potential as both a diagnostic and prognostic biomarker, particularly in TNBC patients.

**SIGNIFICANCE:** An automated and spatially resolved AI tool for sTILs scoring enhances patient risk stratification based on both response to treatment and recurrence-free survival, establishing its relevance as an independent prognostic marker.

## INTRODUCTION

Breast cancer is the second most common cancer worldwide, affecting millions of people each year. Triple negative breast cancer (TNBC), which affects 10-20% of patients, is an aggressive subtype associated with poor prognosis (1–4). Treatment primarily relies on cytotoxic chemotherapy, and options for targeted therapies are limited (1–3). However, TNBC has been found to exhibit greater levels of immunogenicity compared to other breast cancer subtypes (5,6), which presents a unique opportunity to investigate how the molecular and immune interactions within the tumor microenvironment (TME) drive disease progression. Consequently, development of immunotherapies such as pembrolizumab show promise for TNBC treatment (1–3,6–8), but the immune infiltrate is highly diverse in its makeup and magnitude, even within the TNBC molecular subtype. As a result, only a fraction of patients benefit from the addition of immunotherapy (3,7,8), underscoring the need to further our understanding of the underlying biology to overcome the challenges in treating such a heterogeneous disease.

Tumor infiltrating lymphocytes (TILs) have been increasingly acknowledged as a robust predictor of TNBC treatment response with higher TIL densities correlating with improved response to neoadjuvant chemotherapy (3–5,8–11) and patient survival (3–10). Particularly, stromal TILs (sTILs), are a promising prognostic biomarker in breast cancer (9,11), which is typically reported as a fraction of the total stromal compartment area. To standardize the practice of quantifying sTILs, a group of investigators in the International TILs Working Group published a set of guidelines for manual assessment of sTILs in breast cancer (11,12). Although these efforts improved concordance among pathologists, manual scoring remains difficult to standardize due to inter-observer variability, limiting the precision and utility (13,14).

Beyond clinician subjectivity, sTIL assessment is fundamentally challenged by the spatial heterogeneity of lymphocytes themselves. For example, they may form dense, organized structures such as tertiary lymphoid structures (TLS), which are emerging as strong predictors of response (15). Conversely, immune cells may also be dispersed and isolated, hindering signaling events and anti-tumoral activity. Overall, their spatial arrangement across the tumor microenvironment can vary significantly across the tumor microenvironment, altering the prognostic impact of immune infiltration (16–19). While the International TIL Working Group guidelines instruct pathologists to score only sTILs in proximity to invasive cancer as a proxy for capturing functional anti-tumor response, this can be difficult to standardize, is vulnerable to subjectivity, and does not systematically account for local sTIL spatial arrangements (9,12,13,20). Although pathologist assessment of sTILs is currently the accepted practice, the clinical significance of the spatial heterogeneity of sTILs remains largely unclear.

Consequently, there has been an expanding effort to develop automated tools for sTIL quantification using deep learning-based approaches in breast cancer (21–24) and other solid tumors (24–27) to increase reproducibility and assist clinicians in efficiently utilizing TILs as a prognostic marker. Current methods primarily calculate sTIL scores based on the percentage of the stromal area occupied by sTILs, largely ignoring their heterogeneous distribution. Preliminary efforts have been made to spatially map sTILs in H&E-stained images of breast cancer (28,29), implying that local distributions of lymphocytes play an important role in governing a patient’s response to therapy. This is supported by pre-existing biological evidence demonstrating that the proximity between immune subsets and tumor cells in breast cancer is a critical determinant of patient survival (30–32). However, in breast cancer, there still remains a need for tools that move beyond aggregate and density-based measures to quantify the impact of spatial organization of sTILs on its prognostic value while utilizing the entire whole slide image (WSI). Our methods introduce a framework for systematically mapping each sTIL within defined distances from all epithelial cluster boundaries, enabling us to assess how the prognostic value of sTILs may vary based on their precise location with respect to the epithelial cells.

In this paper, we have developed “TILseg”, a biologically intuitive, deep learning-driven pipeline that reproducibly calculates sTIL scores from WSIs of diagnostic biopsies and quantifies the spatial distribution to capture each patient’s inherent immune response, a key insight used to inform treatment planning (33). Our framework provides automated sTIL scoring by sampling the entire WSI (hereafter referred to as global sTIL scores), with the potential to reduce clinician workloads by acting as an assistive tool as well as reducing the subjectivity associated with pathologist expert scoring. We correlate the TILseg generated scores with pathologist scores, as well as compare the predictive power of patient outcomes using computational and manual sTIL scores. We applied our pipeline to two different TNBC cohorts (independently reviewed by several pathologists) and achieved statistically significant stratification of patients based on their recurrence-free survival (RFS) and pathologic complete response (pCR) status using the diagnostic biopsies alone. This suggests that the TILseg sTIL scoring system predicts response to treatment based on the inherent immune response of the patients, making it a potential diagnostic biomarker for risk stratification. Furthermore, we explore how the spatial proximity of sTILs to the epithelial cells in TNBC affects their predictive power with respect to RFS and pCR status, capturing a more complex tumor microenvironment interplay not possible with global scoring. TILseg simultaneously samples the entire WSI and provides spatially constrained sTILs scores around epithelial clusters, improving the prognostic power of TILs for predicting RFS and pCR status.

## MATERIALS AND METHODS

### Data Acquisition and Preparation

In an IRB-approved retrospective study, archived tissue specimens were obtained for a cohort of 60 TNBC patients who previously underwent neoadjuvant chemotherapy (NACT) at our institution. Two patients were excluded from this study due to widespread necrotic regions throughout their tissue biopsies, based on pathologist recommendation. One other patient was lost to follow-up, leaving 57 patients for this study. Tissue sections were cut from formalin-fixed paraffin-embedded (FFPE) diagnostic core needle biopsy specimens obtained prior to treatment. Tissue sections were then stained with hematoxylin and eosin (H&E). All slides were scanned using the Aperio ScanScope AT2 scanner at a microns-per-pixel (MPP) resolution of 0.2525. We will refer to this cohort as the ‘discovery cohort’ hereafter.

A second cohort (independent validation cohort) of 54 TNBC specimens, as well as the associated clinical characteristics and survival data, were collected from our institution in an IRB-approved prospective study. From this cohort, 43 samples were included in the final analysis after excluding patients that were lost to follow-up (n = 9) as well as removing out-of-focus WSIs (n = 1) and widespread regions of active necrosis (n = 1) across the sample set. Tissue sections were cut (4-5 μm), stained with H&E, and then scanned at a MPP resolution of 0.2624. This cohort will be referenced as the ‘independent validation cohort’ hereafter.

From the discovery cohort, 28 WSIs were allocated for training our tissue classifier. The remaining 29 WSIs were split into two internal validation sets: validation I (n = 21), used to iteratively check model performance during training, and validation II (n = 8), which was randomly selected and locked for final, unbiased testing on completely unseen data after model optimization (Table 1).

**Table 1.**
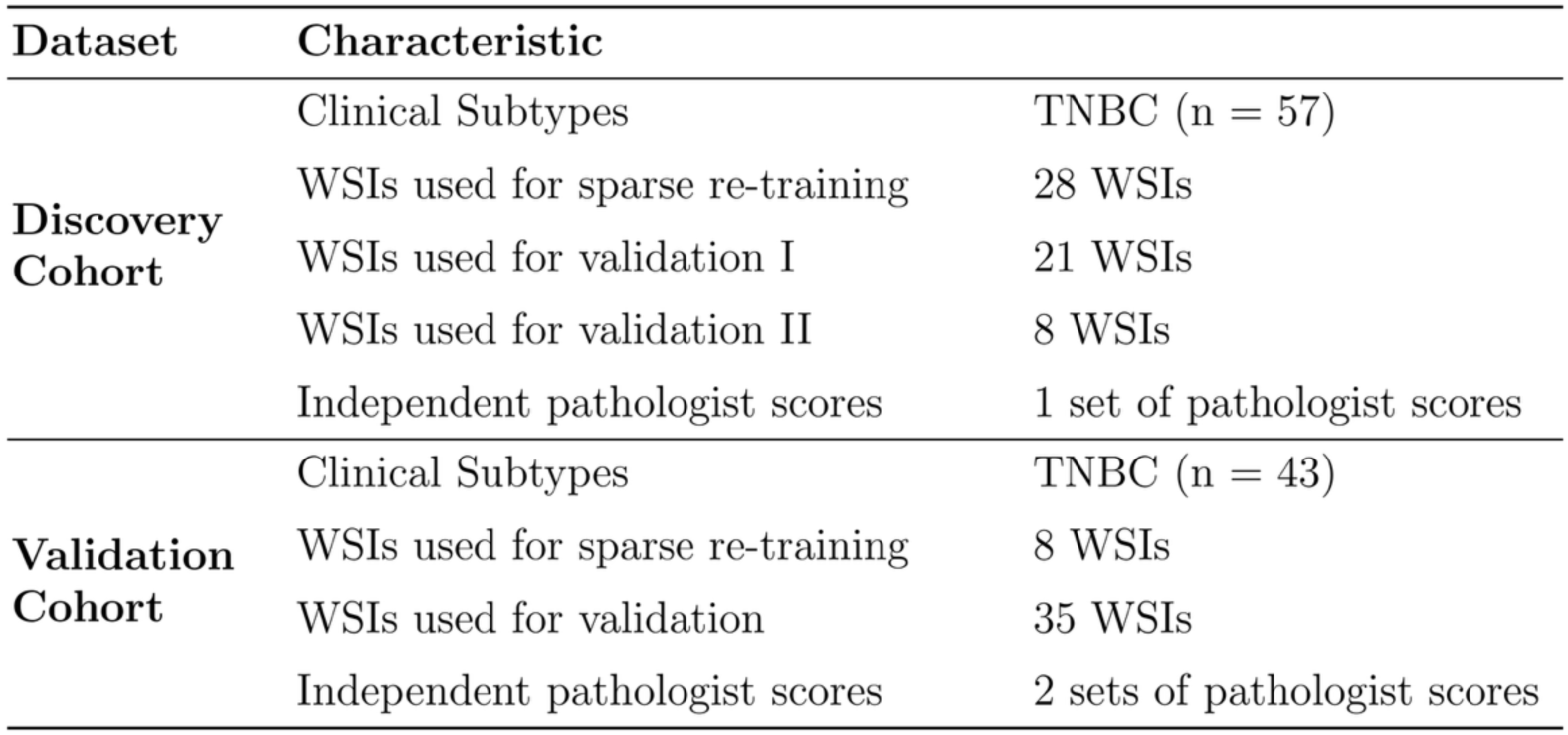
Two independent cohorts of H&E-stained core needle biopsies used in this study.

Using the tissue classifier model trained on the discovery cohort, 8 WSIs from the independent validation cohort were annotated for sparse-retraining. After fine-tuning the model, we qualitatively validated its performance and generalizability on the remaining 35 WSIs (Table 1).

### Manual Pathologist Scoring

In the discovery cohort, a single pathologist assessed areas across each WSI and assigned a single sTIL score for the corresponding patient by estimating the area occupied by immune infiltration as a fraction of the stromal area in areas adjacent to invasive cancer. The pathologist followed guidelines per the International TIL Working Group (12).

In the independent validation cohort, two pathologists discussed and defined standardized guidelines based on the International TIL Working Group by watching scoring videos, practicing on training sets, and reviewing test cases prior to scoring (12). Then, they selected a specific region of interest (ROI) for each patient prior to scoring to ensure that both pathologists are counting the same area and independently assigned an aggregate sTIL score for each patient by estimating the area of immune infiltration as a fraction of the stromal area within the border of invasive tumor for that ROI. Based on their scores, patients were categorized as having either low (0-10%), intermediate (10-40%), or high (40-90%) lymphocyte infiltration. These bins correspond to groups A, B, and C, respectively, per International TIL Working Group guidelines (34). In cases where the pathologists placed the same patient into different groups, the sTIL score was reassessed by both clinicians together to come up with a consensus score. If a patient was assigned different scores between the two pathologists but remained in the same group, the original clinician scores were retained.

### Computational sTIL Scoring Pipeline

For each WSI, the tissue area was selected by annotating around the biopsy specimen region using Aperio Imagescope to exclude unnecessary background. The annotation coordinates were exported as XML files and then used to divide the WSI into 3000 × 4000-pixel patches with 5% padding and 5% overlap, ensuring edge effects are accounted for (Fig. 1A). This patch-based approach allows a deep learning model to efficiently process large medical images. (35) After parsing the WSIs, each patch (50×50 pixels resized to 48×48) was fed as input to the trained tissue classifier (Fig. 1B) to classify stromal, epithelial, and other tissue regions (Fig. 1C). Separately, we applied the StarDist nuclear segmentation model to the same H&E patch in order to identify all nuclei, including both benign and neoplastic, and saved the segmentations as contours (Fig. 1C).

**Figure 1.**
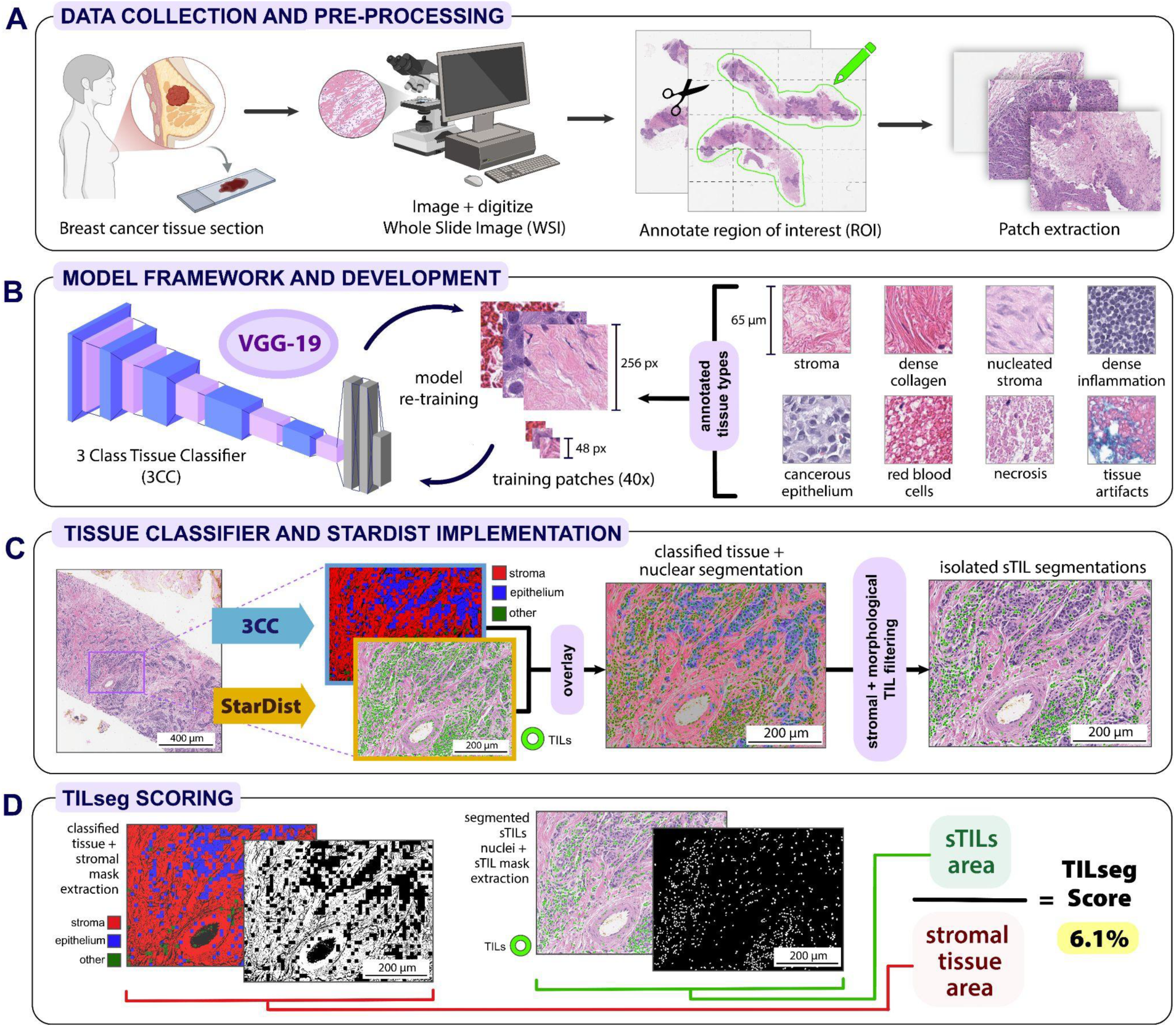
Data preparation, model development, and implementation of TILseg. (**A**) H&E-stained WSIs were collected from two independent cohorts of TNBC patients. The discovery cohort samples were scanned using the Aperio ScanScope AT2 scanner at a resolution of 0.2525 MPP. For each WSI, the tissue area was manually annotated in Aperio Imagescope and then extracted into patches of 3000 × 4000 pixels. (**B**) A pre-trained deep convolutional neural network with a VGG19 model architecture was used to classify H&E-stained images of breast cancer tissue into stromal, epithelial, and other regions (3CC). The model was re-trained using 256 × 256 and 48 × 48-pixel training tiles from WSIs in the discovery cohort. For adaptation to the independent validation cohort, additional WSIs were used to further fine tune the model. The training tiles encompassed a variety of tissue types including loose stroma, dense collagen, nucleated stroma, dense infiltration in the “stroma” class, neoplastic and benign cells in the “epithelium” class, red blood cells, necrosis, and other tissue artifacts in the “other” class. (**C**) Once the WSIs were extracted into 3000 × 4000-pixel patches, model predictions from the 3CC and StarDist were independently implemented in patch-wise manner to classify tissue based on RGB inputs and segment nuclei across the WSI, respectively. Nuclear segmentations outside of the stromal mask or not within the morphological thresholds were eliminated and not considered in the final sTIL score calculation. (**D**) The stromal area was extracted from the 3CC model predictions, while the area occupied by sTILs was derived from the sTIL segmentation contours. The final TILseg score was calculated as the sum area occupied by the sTILs divided by the cumulative sum of the stromal area across all patches.

Since TIL populations in stromal areas have shown to be more prognostic, the stroma mask from the classified image was used to exclude nuclear contours outside of the stroma, effectively eliminating any segmented neoplastic and epithelial cells from our analysis. This was done by eliminating contours that had < 50% overlap with the stromal mask. To further isolate the sTIL population, we applied thresholding specific to the morphology of TILs that differentiate it from larger stromal cells, elongated fibroblasts, and extraneous epithelial nuclei in the stromal region. Since TILs are generally smaller in area and have a more circular shape compared to these other cells, we retained only contours approximately 3-8 μm in diameter as well as a roundness score (defined by Eq. 1) below 1.2 at 0.2525 MPP resolution (with a score of 1.0 indicating perfect roundness). This allows us to exclude large, irregularly shaped, and elongated cells that are typically uncharacteristic of TILs morphology.

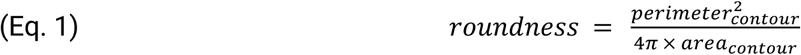

To quantify the extent of stromal infiltration across the entire WSI, we calculated an sTIL score by dividing the summed area of all sTILs by the total stromal area. The total sTIL area was derived by cumulatively adding the areas across all sTIL nuclear segmentations, while the stromal compartment was calculated based on the area of tissue classified as ‘stroma’ by the 3CC. The final sTIL score for the WSI was computed using a formula adapted from the International TIL Working Group Guidelines (Eq. 2).

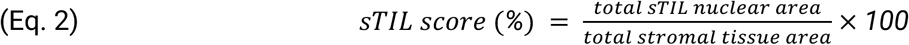

These calculations were carried out on a patch basis, but the total sTIL and stromal areas were summed across all patches to generate a WSI-level sTIL score for each patient (Fig. 1D)

### Model Development and Optimization

In order to segment the breast tissue into three classes (stroma, epithelium, and other), we applied a convolutional neural network (CNN) with a VGG19 model architecture that was previously trained on H&E-stained WSIs of breast cancer (35). The model requires red, green, and blue pixel values as channel inputs to classify an image into three main tissue types: stroma, epithelium, and miscellaneous tissue including necrosis, red blood cells, and other tissue artifacts.

To re-train and tailor the 3-class tissue classifier model (3CC) to our discovery dataset, we selected and annotated 28 cases from the discovery dataset to account for stain and morphological variations. Since the output categories are broad, we further categorized our annotations into more specific local tissue structures present within the H&E-stained images. For the ‘stroma’ class, we differentiated dense collagen fibers, lymphocyte infiltration, areas with lots of stromal cells, as well as desmoplastic collagen fibers to incorporate maximum intra-class heterogeneity to minimize the inter-class confusion, in this case with ‘epithelium’ or ‘others’ class. To encapsulate the different types of epithelial tissue, we annotated normal epithelium, benign pathology, invasive cancer, and ductal carcinoma in situ (DCIS). Finally, areas with necrosis, red blood cells (RBCs), and artifacts such as ink and folded tissue were marked to be incorporated into the ‘others’ class. The annotations were subsequently used to extract two different tile sizes for training, 48×48 and 256×256 pixels, to capture more granular details as well as broader tissue structures, respectively (Fig. 1C). All the tiles were resized to 48×48 pixels prior to entering the 3CC.

The previously trained tissue classifier was fine-tuned to our discovery cohort by initializing with its pretrained weights and unfreezing the last four layers for training using 16,495 ‘stroma’, 11,996 ‘epithelium’, and 8,000 ‘other’ patches. The classes were incorporated with different prior probabilities to approximately reflect the natural abundance of these tissue types in breast cancer and the surrounding TME. We further augmented the data using rotation, shearing, zooming, flipping, brightness and color jitter, as well as shifting. Then, we randomly split the tiles from across the training cohort into three sets: 70% for training, 10% for calibration and hyperparameter tuning, and 20% for testing the model’s generalizability on unseen data. After splitting the data, the last four layers of the model were re-trained over a maximum of 50 epochs using the Adam optimizer with a learning rate of 0.001, with a decay rate of 0.1 if there was a plateau in the validation accuracy for five consecutive epochs. The training was stopped early if there was no improvement in the validation accuracy for ten consecutive epochs, and that iteration of the model is saved as the optimal checkpoint.

In order to adapt the model we trained on the discovery cohort to the staining and imaging variations in the independent validation cohort, we further carried out sparse retraining by annotating regions of interest from a small number of WSIs (n = 8) to parse into stroma, epithelium, and other tiles for transfer learning. Following a similar workflow as the discovery cohort, the tiles across the eight patients were split into 70% for training, 10% for calibration, and 20% for validation. In this case, only the output layer of the previous model was subsequently fine-tuned under identical training constraints (e.g., early stopping, learning rate decay) to ensure consistency in performance and evaluation across cohorts.

### StarDist Nuclear Segmentation

In order to segment all nuclear structures, we utilized a StarDist model, *2D_versatile_he* (36), pre-trained on publicly available H&E image datasets to perform instance segmentation of star-convex objects. Specifically, the model we implemented uses RGB inputs to identify nuclei in H&E-stained histopathology images. We selected this model for its computational efficiency compared to other pixel-wise segmentation models as well as its ability to separate overlapping nuclei specific to brightfield H&E-stained images. Since the pretrained model was performing accurately and reliably on our images, no additional re-training was carried out.

### Scoring sTILs at Varying Distances to Epithelial Clusters

The TILseg pipeline outputs 3-class tissue classification masks, with pixel-wise segmentation of “epithelium” and “stroma” regions, as well as sTIL masks for each patch of a WSI. To analyze the spatial context of the digitally segmented sTILs and their prognostic value, we first stitched the patches together to reconstruct the 3CC output of the contiguous WSI (Fig. 2A). Then, we calculated a patient sTIL score (Eq. 2) from a spatially defined subset of stroma across the stitched WSI by selecting a population of sTILs for scoring based on two tunable parameters: 1) the maximum distance around an epithelial cluster that stroma will be scored and 2) the minimum size of an epithelial cluster for which to draw that distance.

**Figure 2.**
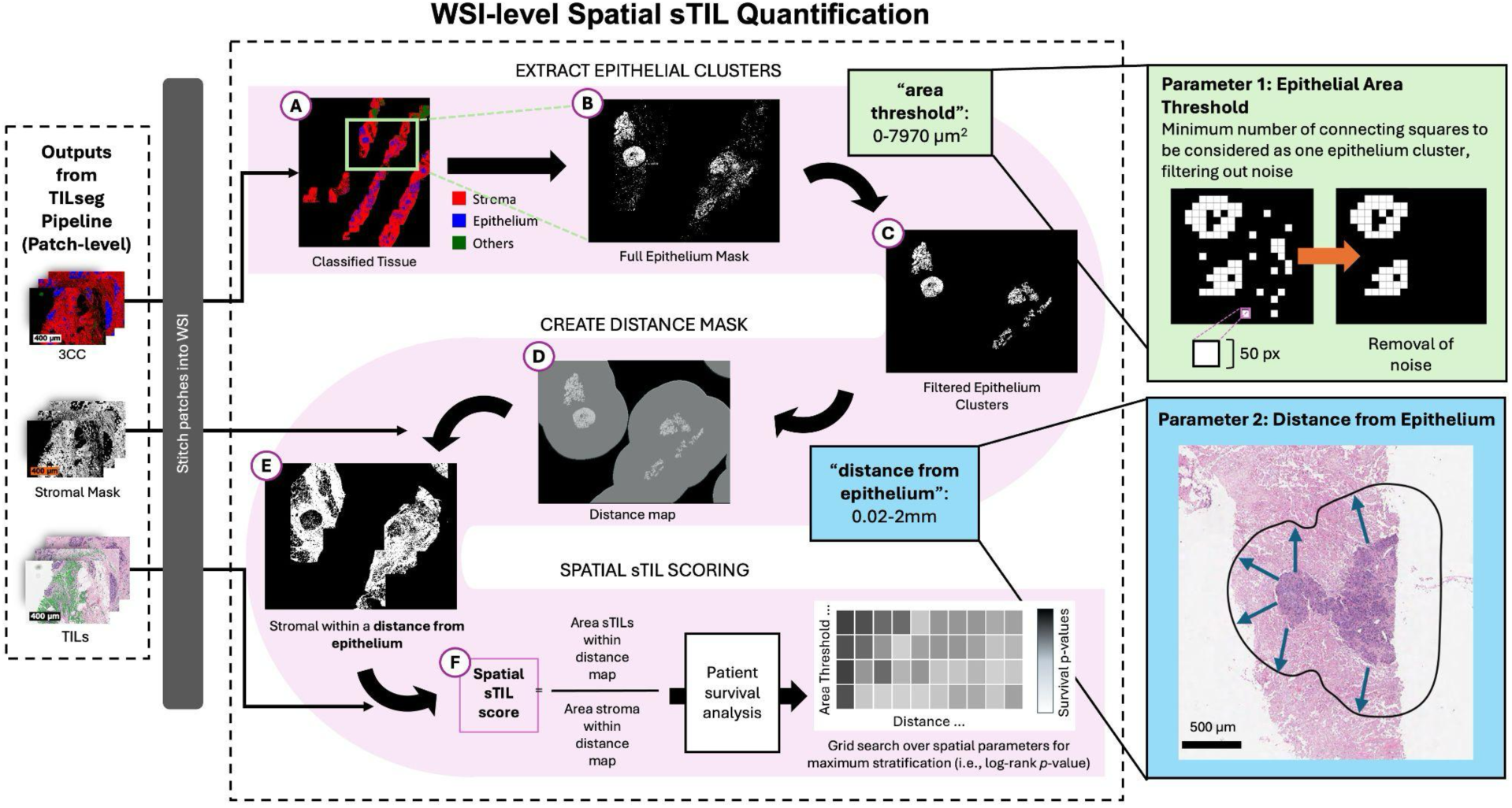
Analysis workflow of sTILs in spatial subsets of stroma across a WSI. **(A)** TILseg 3CC outputs on the patch-level were stitched to form the contiguous WSI. **(B)** Binary mask of epithelia generated from 3CC WSI tissue predictions. **(C)** The “epithelial cluster area threshold” parameter is applied to remove small, noisy epithelial cluster predictions from 3CC. **(D)** The “maximum distance from epithelial cluster” parameter is applied to create a mask of regions that are within the specified distance around epithelial clusters. **(E)** The distance mask is multiplied with the stroma mask from 3CC output (A) to generate a binary mask of stroma regions that are within the required distance from epithelial clusters. **(F)** sTIL score is computed for this spatial subset of stroma computed in (B)-(E). This process is repeated for multiple combinations of “epithelial cluster area threshold” (C) and “maximum distance from epithelial cluster” (D) parameters to generate patient-level sTILs scores for different spatial subsets of stroma across the slide. Survival analyses (*i.e.*, training a Cox regression model) are performed to identify the spatial stroma subset whose sTIL scoring yields the highest association with TNBC patient recurrence-free survival after neoadjuvant chemotherapy.

The second parameter in this “spatial” sTIL score is the minimum size of regions classified as epithelium by the 3-class tissue classifier of the TILseg pipeline (Fig. 2B), which must be met for a region to be included as an “epithelium cluster” in this calculation (Fig. 2C). To filter out noise from erroneous predictions of small epithelial regions from 3CC, we disregard regions of epithelia that have a smaller area than the “minimum epithelial cluster size” parameter (Fig. 2C). Next, we generate a distance mask of regions that are within the “maximum distance from epithelial cluster” parameter (Fig. 2D). This distance mask is multiplied with the stroma mask from the 3CC output to identify regions of stroma that are within the specified distance of sufficiently large epithelial clusters (Fig. 2E). We examined distances of 0.02-2.0 mm from “epithelium clusters”. We investigated minimum epithelium cluster areas ranging from 0 (considering all epithelial regions predicted)-7970 µm^2^ (Fig. 2) to filter out regions of small, noisy epithelia that would confound the spatial scoring of sTILs. The area of the spatially constrained stroma mask is the denominator in the spatial sTIL score (Eq. 2). Additionally, sTILs whose centroids fall outside of the spatially constrained stroma mask are excluded, and the area of the remaining sTILs is the numerator in the spatial sTIL score (Fig. 2F, Eq. 2). Spatial sTIL scores were computed for each combination of “minimum epithelium cluster area” and “maximum sTIL distance from epithelium cluster” parameters explored (Supp. Fig. 3).

**Figure 3.**
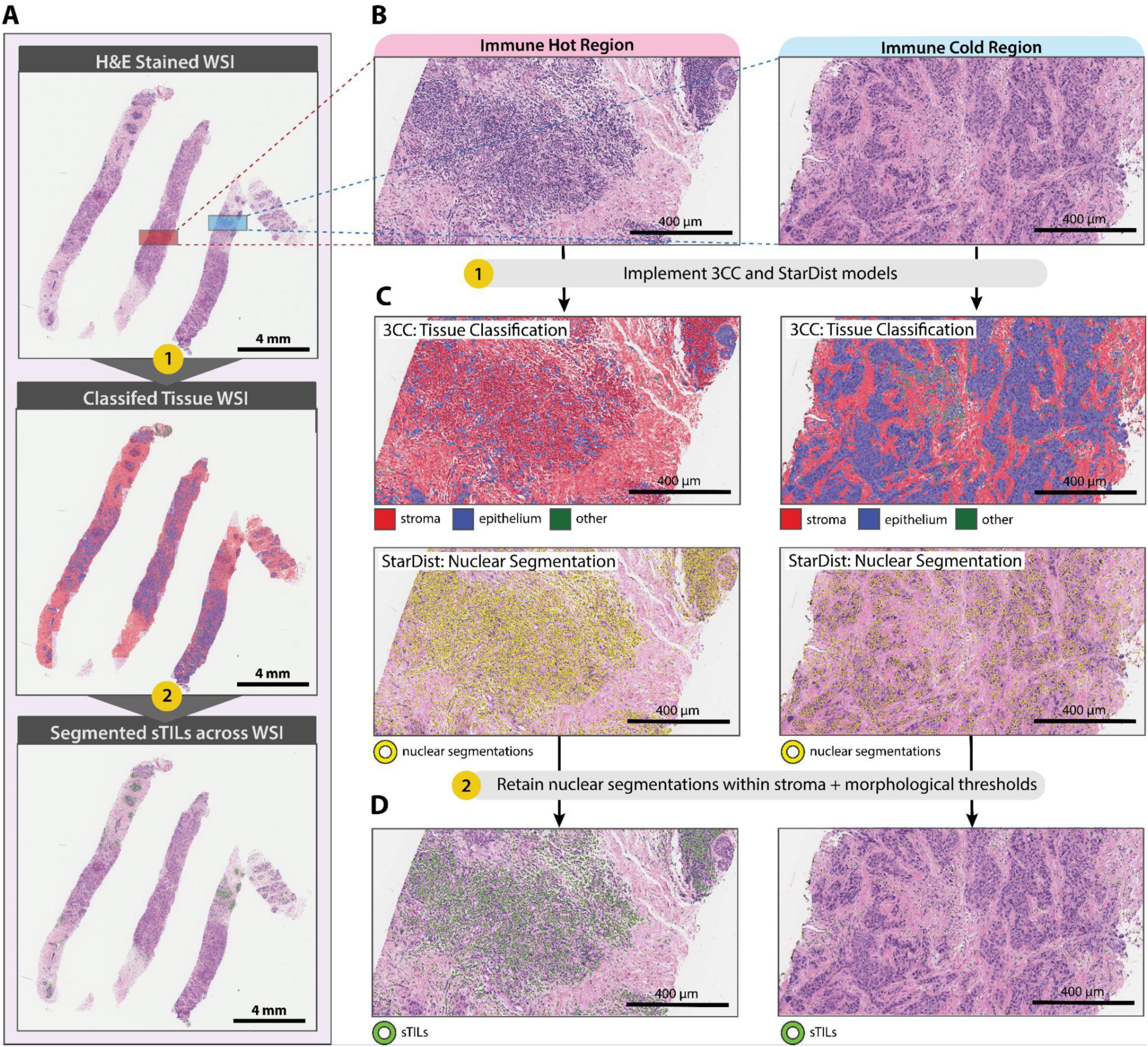
Visual overlay of TILseg outputs on two representative regions. (**A**) TILseg comprehensively processes and visualizes H&E staining (top), tissue classification (middle), and sTIL segmentation (bottom) at the WSI-level with 0.2525 MPP resolution. (**B**) Analysis of H&E-stained core biopsy including regions with high (left) and low (right) immune infiltration across the WSI. (**C**) Corresponding visualization of 3CC tissue segmentation of the biopsy core into stromal (red), epithelial (blue), and other (dark green) regions overlaid on the H&E image. Separately, StarDist was used to segment all nuclei across the entire WSI (yellow). Each WSI was analyzed in a patch-wise manner, but the outputs were subsequently stitched together to visualize the whole slide. (**D**) sTIL segmentation (light green) across the biopsy. Segmentations of benign and neoplastic cells were excluded from the final visualization of sTILs.

### Clinical Outcomes

Clinical outcomes in the discovery cohort (n = 57) were also defined by the duration of RFS, which was measured from the date of surgery until the last documented clinical follow-up or until the recurrence of the tumor after surgery or death. The Mann-Whitney *U* test was used to assess the difference in TILseg scores between good and poor outcome groups. (37) Five patients who did not have a recurrence at the last clinical follow-up but had a follow-up time of less than 3 years post-surgery were omitted from this analysis (n = 51). Patients who had a recurrence within 3 years of surgery (n = 12) were considered a poor outcome, while those who did not (n = 39) were the good outcome group. We also assessed the difference in pCR status between the two outcome groups using Fisher’s Exact test. One patient did not have pCR status evaluated and was not used for this analysis (n = 50).

We defined pCR as no residual tumor (ypT0) and no lymph node involvement (ypN0) post-neoadjuvant chemotherapy in our analysis. One patient who did not have a pCR status evaluated in the discovery cohort was omitted from this analysis (n = 55 patients, n = 15 pCR, n = 40 non-pCR). The Mann-Whitney *U* test was used to assess the significance of the difference in TILseg scores between the pCR and non-pCR groups.

### Statistical Analysis

To assess the strength of correlation between computational sTIL scores generated by TILseg and manual pathologists’ scores, we calculated the Spearman’s rank correlation coefficient (38) in both the discovery (n = 55) and independent validation (n = 43) cohorts. Two patients in the overall discovery cohort (n = 57) did not have manual pathologist scores, so they were not included in this analysis.

Further, we assessed the strength of association between TILseg scores, as a continuous predictor, and RFS in the discovery cohort (n = 57) using multivariate Cox regression analyses, adjusting for tumor stage at the time of diagnosis. Influence diagnostics for the Cox model were conducted using DFBETAS analysis (standardized differences in regression coefficients) (39). This method re-estimates regression coefficients after leaving out one observation at a time, similar to leave-one-out cross-validation or the jackknife, and calculates the change in the regression coefficients, normalized by the standard error, relative to when all observations were used. This can identify highly influential observations (e.g., an outlier) that disproportionately impact model estimates in an objective manner. A threshold of 0.56 standard errors was used to identify influential observations, based on a guideline of two divided by the square root of the number of events (n = 13) (Supp. Fig. 4A) (39).

**Figure 4.**
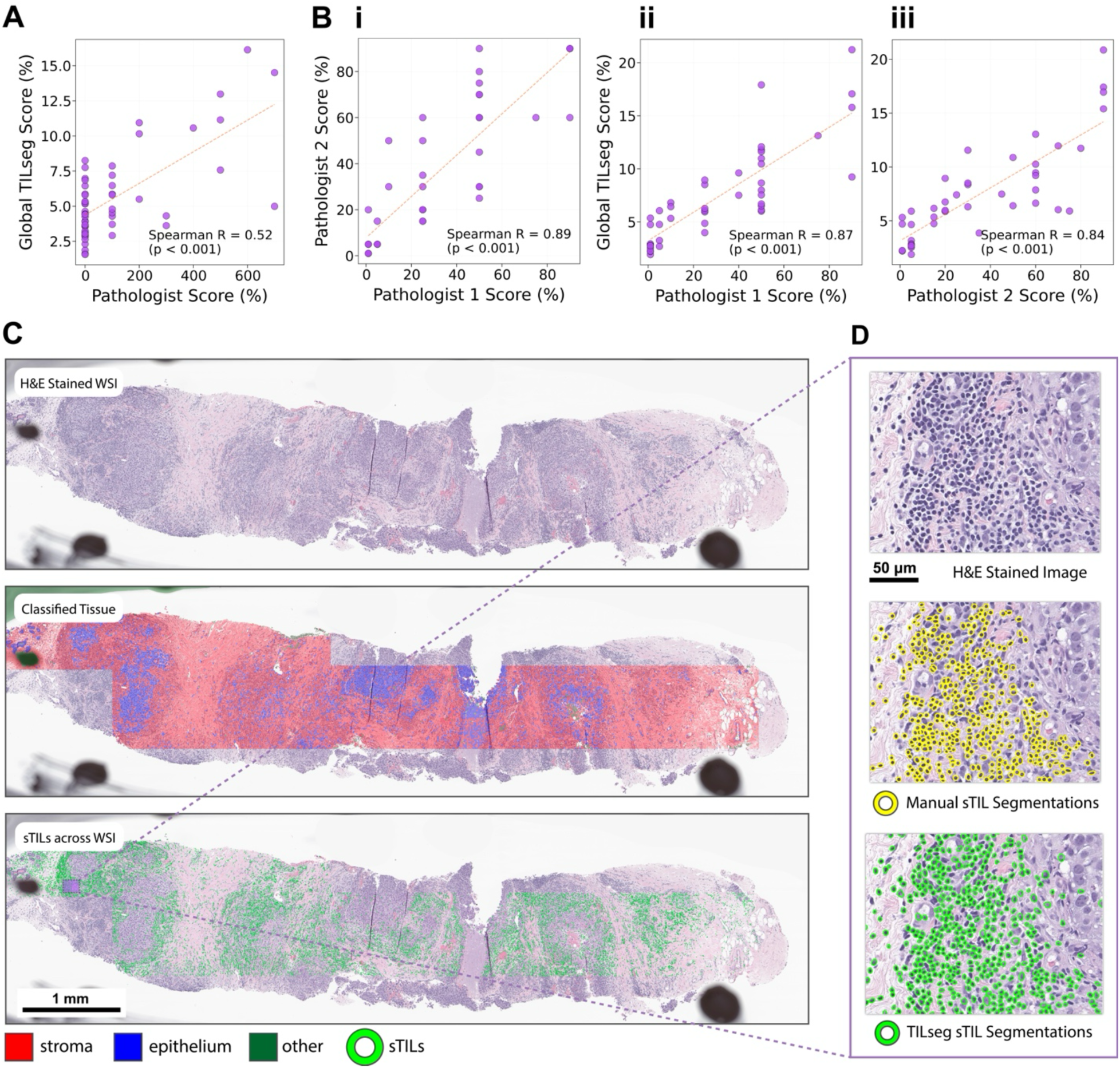
Comparison of computational TILseg sTIL scores with manual pathologist scores. (**A**) Correlation between manual sTIL scores and TILseg scores in the discovery cohort (n = 55) where each point represents a single patient (Spearman R = 0.52). Visual scores were obtained from one pathologist following guidelines from the International TILs Working Group, while the entire biopsy core was processed through TILseg to obtain global sTIL scores. (**B**) (**i**) Inter-pathologist agreement for sTIL assessment (Spearman R = 0.89) in the independent validation cohort (n = 43). Prior to scoring, two pathologists selected a single region of interest (ROI) and agreed upon standardized guidelines adapted from the International TILs Working Group. (**ii, iii**) Spearman correlation between manual sTIL scores and TILseg scores in the independent validation cohort (Spearman R = 0.87, 0.84). After independent scoring, the two pathologists agreed upon consensus scores if their original assessments had placed patients into different clinical categories. Otherwise, their scores remained the same. Each pathologist’s scores, after consensus scoring, were compared with TILseg scores. (**C**) Performance of TILseg on a WSI from the independent validation cohort. The pipeline is able to adapt to variations in staining after fine-tuning the 3CC model. (**D**) Comparison between manually segmented sTILs (yellow) and TILseg sTIL segmentations (green).

In regards to the discovery cohort, DFBETAS analysis (39) of the Cox regression coefficient of the global TILseg scores resulted in one exceptionally influential patient (Supp. Fig. 4A), which was excluded from subsequent statistical analyses (n = 56). Analyses including this patient included (n = 57) are presented in Supp. Figs. 4 and 5. While the data point disproportionately affected the Cox regression results, the conclusions from the other statistical tests remain relatively unaffected by the omission of this patient.

**Figure 5.**
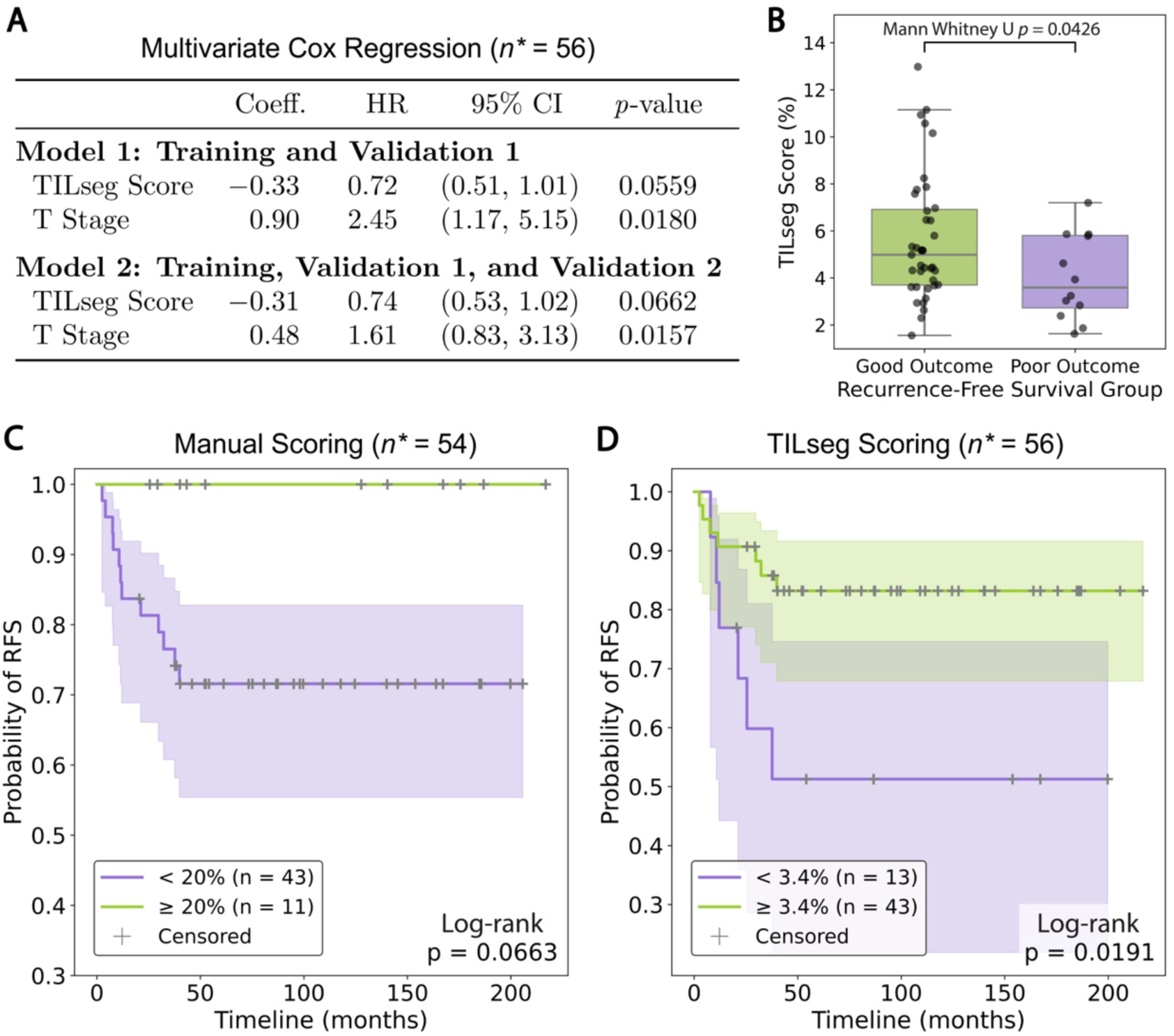
Global TILseg scoring as a predictive biomarker for recurrence risk post-neoadjuvant therapy in the discovery cohort. (**A**) A multivariate Cox regression model was fitted on WSIs in the training and validation I subsets (n = 48), showing the TILseg score is an independent prognostic factor with a 28% reduction in recurrence hazard (HR = 0.72 per 1% increase, 95% CI [0.51-1.01], p = 0.0559) after adjusting for clinical tumor stage (HR = 0.90 per stage increase, 95% CI [1.17-5.15], p = 0.0180). A second multivariate Cox regression model was fitted on WSIs in the training, validation I, and validation II samples (n = 56). Consistent directional effects were observed where TILseg remained associated with improved RFS (HR = 0.74 per 1% increase, 95% CI [0.53-1.02], p = 0.0662), independent of tumor stage at diagnosis (HR = 0.48 per stage increase, 95% CI [0.83-3.13], p = 0.0157). The 8 patient WSIs in the validation II dataset were not used for any part of the TILseg pipeline development, and these patient TILseg scores resulted in an association with RFS that remained strong. (**B**) Patients in the discovery cohort who recurred within 3 years (poor outcome, n = 12) had significantly lower TILseg scores than patients who did not recur within 3 years (good outcome, n = 39). Kaplan-Meier stratification of RFS is shown for the discovery cohort by (**C**) manual scoring (n = 54) and (**D**) TILseg scoring (n = 56). A threshold of ≥ 20% sTILs was used for manual assessment while a cutoff of ≥ 3.4% was used to stratify patients based on the optimal log-rank *p*-values. * indicates that one highly influential and unrepresentative patient was excluded from this figure and is shown in Supp. Fig. 4.

Then, we used Kaplan-Meier (KM) analysis with the log-rank test to visualize and assess the degree of stratification in the discovery cohort into low- and high-risk groups with respect to RFS based on their sTIL scores. For both manual pathologist scoring (n = 54) and computational scoring (n = 56) with TILseg, the sTIL score threshold was determined by using the optimal cutoff that yielded the lowest log-rank p-value in the discovery cohort, with at least 10 patients above and below the threshold. We also assessed the degree of stratification of RFS achieved by the established biomarker of pCR status following NACT as a comparison, using a KM curve and log-rank test (n = 55). In the independent validation cohort, the association between TILseg scores and pCR status was assessed for the subset of patients who received NACT (n=30 patients, n=15 pCR, n= 15 non-pCR). However, the association between TILseg scores and RFS outcomes could not be assessed due to the lack of long-term follow-up data in this cohort.

To identify the optimal spatially defined sTILs population for prognostic stratification, we conducted a systematic evaluation of sTIL quantification across multiple spatial compartments defined by epithelial area thresholds and distance from tumor epithelium. We utilized multivariate Cox regression modeling to test the association (independent of tumor stage at diagnosis) between spatial TILseg scoring and RFS across a range of spatial parameters.

## RESULTS

### Patient Characteristics

The discovery cohort comprised 57 TNBC patients who underwent NACT treatment. The tumor stage distribution of the samples used in our study from the discovery cohort was as follows: stage 1 (16%, n = 9), stage 2 (42%, n = 24), stage 3 (37%, n = 21), and stage 4 (5%, n = 3). At a median follow-up of 5.6 years (range from 0.2 to 16.6 years), 75% of patients (n = 43) remained recurrence-free.

The independent validation cohort comprised 43 TNBC patients who were independently scored by two pathologists. These patients had primarily early-stage tumors (stage 1-2) with 30% of them at stage 1 (n = 13), 51% (n = 22) at stage 2, 14% (n = 6) at stage 3, and 5% (n = 2) who were not assigned any clinical stage. The overall median follow-up time after surgery was 2.2 years (range from 0.2 to 3.8 years).

### TILseg: a deep-learning framework for sTILs segmentation and scoring with whole-slide sampling

TILseg quantifies sTILs across the entirety of the extracted tissue area to comprehensively and systematically analyze immune infiltration (Fig. 3). We exhaustively profile the sTIL distribution across each WSI with minimal manual inputs, capturing spatial heterogeneity in immune infiltration that may be missed by traditional pathologist assessments.

The workflow integrates two deep learning models in a sequential manner: first the 3CC model segments the entire tissue into stromal, epithelial and other compartments to capture the overall tissue architecture and second StarDist model performs nuclear segmentation to identify individual cells across the WSI. This hierarchical approach enables precise localization of immune cells within their relevant tissue microenvironments. Fig. 3A shows the outputs of running a representative H&E-stained section of a biopsy from our discovery cohort through the TILseg pipeline, including the identification of stromal and epithelial regions and segmentation of sTILs across the WSI. Fig. 3B shows two zoomed-in regions of high and low infiltration from the section. Fig. 3C shows the accurate prediction of the 3CC on the zoomed-in regions into epithelium and stroma, along with nuclear segmentation predictions using a pre-trained StarDist model. To specifically identify sTILs, we implemented a two-stage filtering where nuclear segmentations were first spatially constrained to stromal compartments as defined by the 3CC classifier (Fig. 3C). Remaining nuclei are additionally filtered by morphological thresholding based on area and roundness (Fig. 3C) to remove larger stromal cells, extraneous epithelial nuclei, and elongated fibroblasts from sTIL scoring. This way, we remain with isolated sTILs for patient-level scoring. This integrated pipeline reveals spatial heterogeneity in immune cell distribution demonstrating dense lymphocytic infiltration in the immune hot region and sparse immune cell presence in the immune cold region. This automated approach enables objective, reproducible, and spatially resolved quantification of sTILs across entire tissue sections, providing comprehensive spatial immune profiling data for downstream prognostic and predictive modeling in breast cancer.

### Comparison of TILseg scoring and three pathologists across two independent data cohorts

To validate the accuracy and clinical utility of the TILseg sTILs scoring, we performed comprehensive comparisons against expert pathologist assessments. TILseg derived global sTIL scores in the discovery cohort (n = 57) demonstrated moderate concordance with pathologist assessments (Spearman R = 0.52, *p* < 0.001) (Fig. 4A), capturing the overall immune landscape while maintaining consistency across low and high lymphocytic infiltration levels. While the entire WSI for each patient was scored by TILseg, the areas scored by the pathologist were unmarked and sampling differences likely explains in part the moderate concordance in scoring.

To further evaluate performance, we conducted a detailed inter-rater reliability analysis comparing TILseg against two independent expert breast pathologists who manually scored sTILs in identical regions of interest. Since the ROIs were clearly defined for each patient in the independent validation cohort (n = 43), we restricted our analysis to these regions for consistent TILseg scoring and comparison with manual scores. The manual scores from these marked ROIs showed high inter-pathologist agreement between two clinicians, yielding a Spearman correlation coefficient of R = 0.89 (*p* < 0.001) prior to consensus scoring (Fig. 4B, i). The same ROIs were subsequently scored by TILseg and then compared with Pathologist 1 and Pathologist 2 using their consensus scores (if the initial scores by the two pathologists differed significantly). In contrast to the discovery cohort, there was a significant improvement in agreement between TILseg and two pathologists in the independent validation cohort. Respectively, this yielded correlation coefficients of R = 0.87 (*p* < 0.001) and R = 0.84 (*p* < 0.001) (Fig. 4B, ii-iii), approaching inter-pathologist agreement (with standardized assessment guidelines) for quantifying lymphocyte infiltration in the same areas. This illustrates the importance of standardized guidelines for scoring TILs, as it increases the inter-pathologist concordance and shows the reliability of the TILseg scores in computationally scoring TILs. Overall, this indicates that TILseg and pathologists consistently rank patients similarly in terms of TIL infiltration, despite a difference in scoring scales/ranges.

The consistently high correlations across all comparisons indicate that TILseg captures biologically relevant variation in immune infiltration with reliability compared to expert manual assessment. Fig. 4C illustrates the results of the complete pipeline on a representative breast cancer core biopsy from the independent validation cohort with tissue classification revealing heterogeneous stromal (red) and epithelial (blue) compartments and the final sTIL segmentation map showing the spatial distribution of immune cells (green) across the tissue. Fig. 4D provides a detailed comparison of the manual (yellow circles) versus automated sTIL detection (green circles) at high magnification, demonstrating TILseg’s ability to accurately identify individual lymphocytes based on their characteristic morphology. The high degree of spatial overlap between manual and automated segmentations confirms that TILseg successfully carries out expert-level TILs identification at single-cell resolution.

### Automated scoring of sTILs in diagnostic core needle biopsies stratify recurrence risk in TNBC

To evaluate the prognostic value of the automated sTIL quantification, we performed survival analysis in the discovery cohort with long-term clinical follow-up data (median 5.6 years, range from 0.2 to 16.6 years). The independent validation cohort was excluded from this analysis due to insufficient follow-up data. A multivariate Cox regression model was fit for patients in the training and validation I subsets of the discovery cohort (n = 48) where TILseg was found to be an independent prognostic factor for recurrence free survival (HR = 0.72, 95% CI [0.51-1.01], p = 0.0559) after adjusting for clinical tumor stage at the time of diagnosis (Fig. 5A), with higher TILseg scores being associated with improved outcomes. After incorporating the scores from patients in validation II, which were not used for training or evaluation during model development (n = 8), the p-value remained trending towards significance (HR = 0.74, 95% CI [0.53-1.02], p = 0.0662), affirming the robustness and predictive power of TILseg scoring. The hazard ratios below 1.0 for TILseg scores indicate that higher stromal immune infiltration is associated with reduced risk of recurrence, consistent with established biological understanding of anti-tumor immune responses in breast cancer. (3–10)

Fig. 5B further illustrates the clinical relevance of TILseg scoring by comparing the scores between patients stratified by long-term outcomes. Patients with good outcomes (no recurrence within 3 years, shown in green) demonstrated significantly higher median TILseg scores (∼5%) compared to patients with poor outcomes (recurrence or death, shown in purple) with median TIL scores (∼3.5%) with a statistically significant Mann-Whitney U (p-value = 0.0426).

To further assess the discriminatory power of TILseg scores, we carried out Kaplan-Meier survival analysis comparing patients stratified by immune infiltration levels. Manual scoring by a single pathologist trended towards but did not reach statistical significance for stratifying patients by RFS (log-rank *p* = 0.0663) using a cutoff of 20% (Fig. 5C). TILseg improved the prognostic stratification of high vs low-risk patients (log-rank *p* = 0.0191, Fig. 5D) using a cutoff of 3.4%. The cutoff thresholds for both manual pathologist and computational TILseg scoring were chosen based on the optimal log-rank *p*-value. Patients with TILseg scores > 3.4% exhibited significantly better RFS with ∼83% probability of remaining recurrence free at 16.6 years, compared to only ∼53% for patients with scores < 3.4% at 15.3 years. These findings suggest that the TILseg scoring can act as a promising biomarker for risk stratification in breast cancer.

### Evaluating the prognostic role of spatially-resolved sTILs around epithelia

As the “maximum distance from an epithelial cluster” parameter varies, the TILseg score changes to reflect shifts in immune cell density across the tissue microenvironment (Fig. 6A) as seen in regions 1 and 2 which indicate areas selected for detailed examination. It is evident from Fig. 6B (region 1) and 6C (region 2) that different sTIL spatial distributions are being sampled by incorporating the spatial TILseg scoring system. The left panel shows the original H&E image with sTILs marked in green, the middle panel illustrates the 100 microns distance threshold, and the right panel demonstrates the spatial TILs distribution at the 50 microns threshold, focusing specifically on lymphocytes in the immediate peritumoral microenvironment. Region 1 reveals substantial lymphocytic infiltration in both the peritumoral and distant stromal areas, whereas region 2 displays a different spatial pattern characterized by more compact tumor architecture, lesser intervening stroma and scattered lymphocytes primarily concentrated at the invasive front. The distance-based filtering (middle and right panels) reveals how different spatial thresholds capture distinct immunological niches: the 100 μm threshold encompasses both the invasive boundary and shallow penetration into surrounding stroma, while the 50 μm threshold isolates lymphocytes at the immediate tumor-stroma interface. We have illustrated both a qualitative and quantitative statistical impact of spatially resolved sTILs scoring on patient assessment and stratification. This spatially resolved approach addresses a key limitation of global scoring by distinguishing between different zones of immune infiltration, with immune cells closer to tumor boundaries representing active immune surveillance while distant immune infiltration representing background immune activity. This would provide a foundation for investigating whether proximity-based immune metrics offer enhanced prognostic or predictive value.

**Figure 6.**
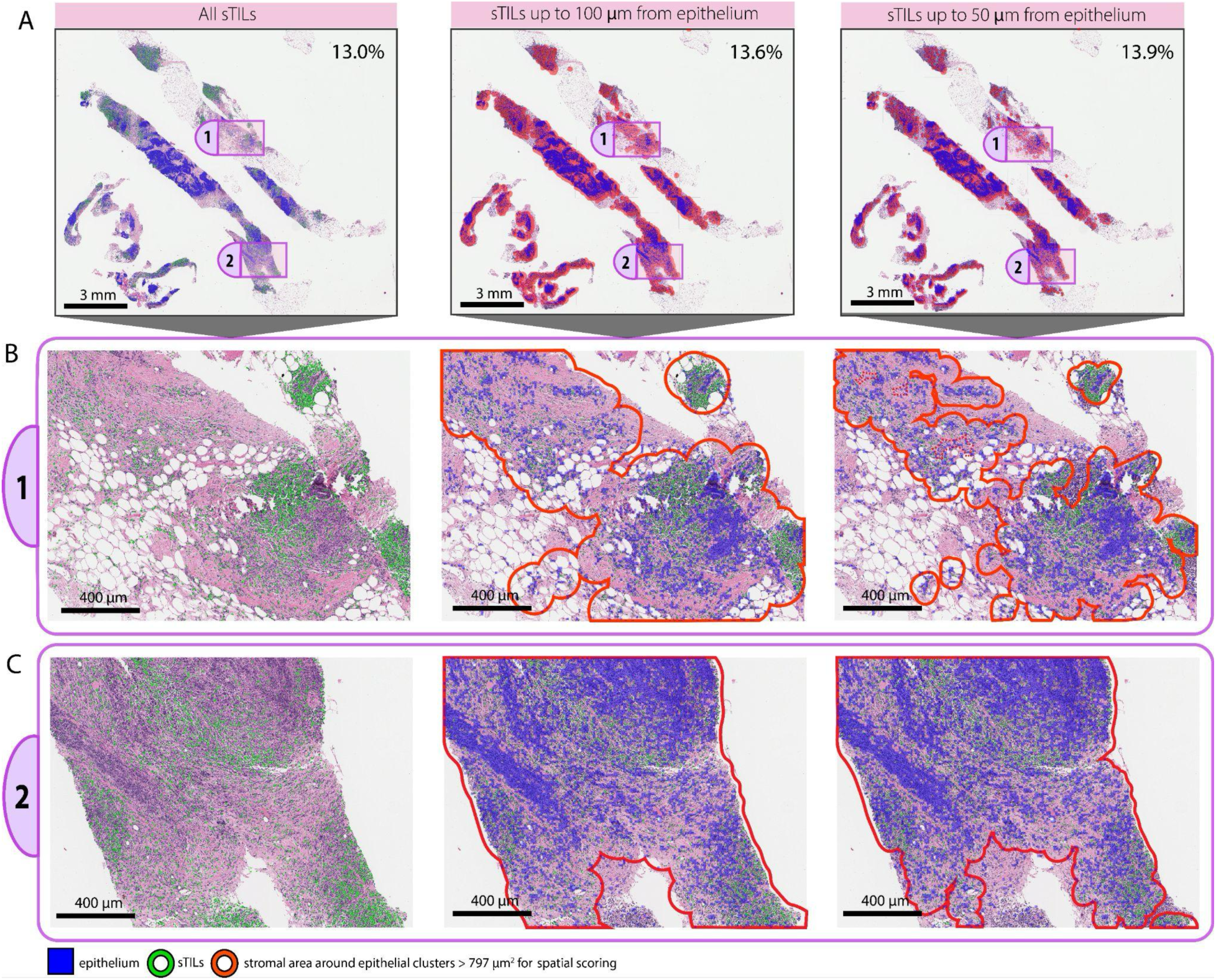
Visualization of TILseg scoring in spatial subsets of a WSI. (**A**) The 3CC identifies epithelial tissue as well as the constrained stromal region at 100 and 50 μm from epithelial clusters > 797 μm^2^ in area across the entire WSI. While the global score is 13.0% sTILs when scored by TILseg, the spatial TILseg score increases to 13.6% and 13.9% sTILs as the total stromal region incrementally decreases to consider only regions 100 and 50 μm from epithelial clusters, respectively. (**B**) Region 1 and (**C**) region 2 illustrate the spatial heterogeneity of sTILs surrounding invasive carcinoma across the breast cancer TME. In these areas, TILseg is able to identify sTILs within 100 μm and 50 μm of epithelial clusters > 797 μm^2^ in area.

Scoring stroma within a large distance around epithelial clusters (*e.g.*, 0.5-2.0 mm) showed relatively weaker association with RFS, as illustrated by the significance (*p-*value) displayed in the heatmap (Fig. 7A). It is important to note that even though TILs up to about 0.5 mm are prognostic towards predicting RFS, the contribution of TILs at closer distances from the epithelial clusters (< 50 µm) maximize its overall predictive power. The biological hypothesis is that these lymphocytes play a crucial role in the patient’s inherent immune response against the progression of breast cancer and other disease types. (30,40,41) However, at very small distances (< 20 µm), we would ignore too many sTILs that play a significant role in the immune response against the disease. At farther distances (0.5-2.0 mm), less prognostic TILs are incorporated, resulting in higher p-values. From our iterative analysis (Fig. 7A), we found that scoring sTILs within 50 µm of epithelia yielded the strongest association with a future TNBC recurrence after neoadjuvant chemotherapy and surgery.

**Figure 7.**
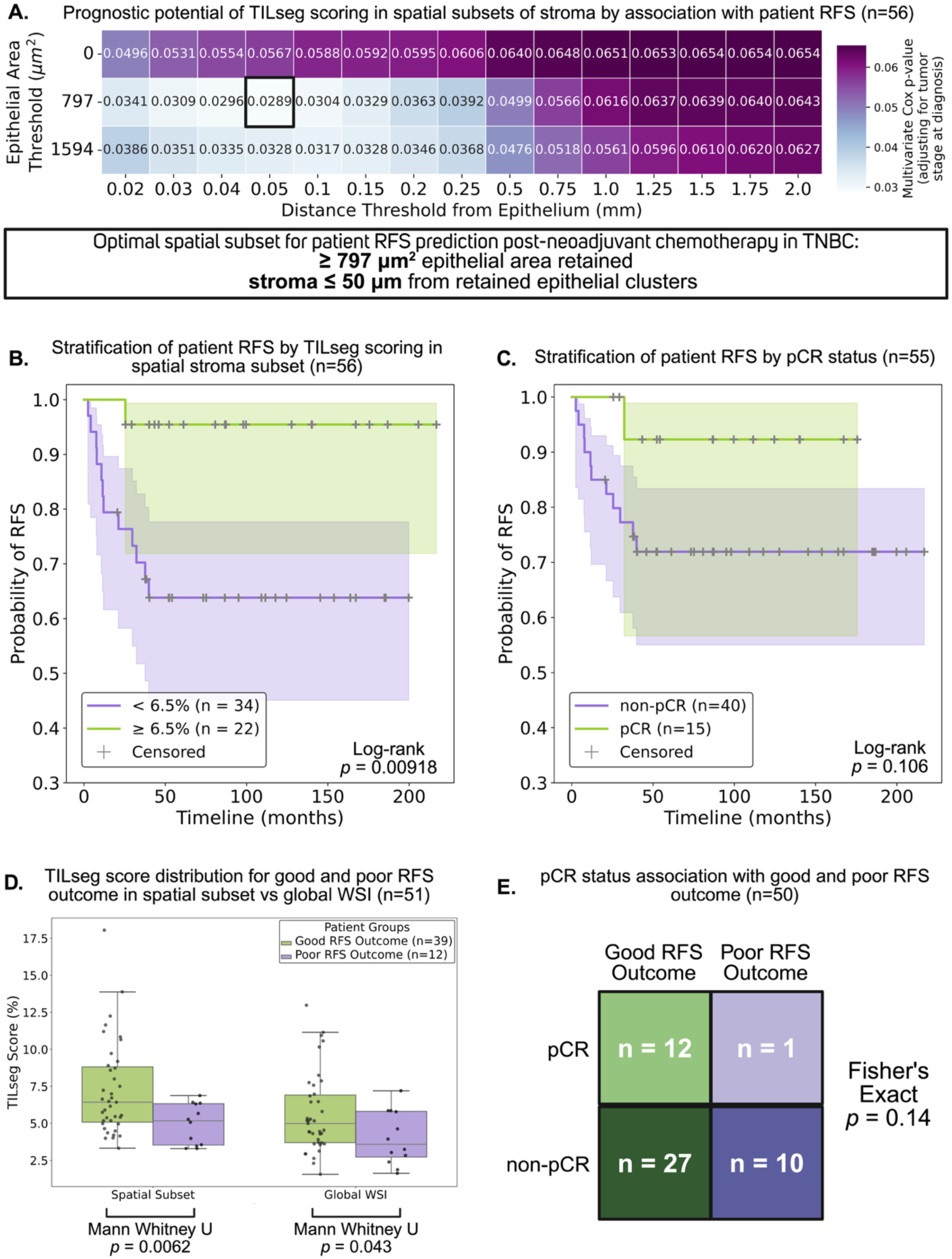
TILseg scoring in a spatial subset of diagnostic biopsies as a predictive biomarker for recurrence outcome in the discovery cohort. **(A)** Multivariate Cox proportional hazards regression *p* values for continuous spatial TILseg scoring in different subsets of stroma around epithelia using diagnostic biopsies from the discovery cohort (n = 56 TNBC patients). The Cox model accounts for tumor stage at diagnosis. The heatmap shows the change in the predictive power of TILs with increasing stromal distance from the epithelial clusters. The columns denote the distance from epithelial clusters within which stroma was scored for sTILs and the rows indicate the size below which epithelial clusters were considered as noise and removed from the scoring analysis. On the y-axis, 0, 797, and 1594 µm^2^ correspond to 0, 5, and 10 kernels (50 x 50 pixel) of 3CC predictions, respectively. A lower *p* value in the heatmap illustrates a more significant association of the spatial subset TILseg score with patient RFS independent of the tumor stage at diagnosis. The spatial subset of stroma with the lowest *p* value (most significant TILseg score association with RFS) is highlighted within a black box. **(B)** Kaplan-Meier curve shows significant improvement in the stratification of patient RFS by spatial TILseg scoring using the suggested optimal spatial stromal pocket and epithelial cluster size (log-rank test). A threshold of ≥ 6.5% sTILs was used as the cutoff based on the optimal log-rank *p*-value. (**C**) Patient pCR status has a much weaker stratification potential of RFS. **(D)** TILseg scoring in a spatial subset of stroma within 50 µm of epithelial clusters that are at least 797 µm^2^ results in a significantly better stratified distribution of the scores (Mann Whitney U *p* = 0.0062) across Good and Poor RFS patient groups than scoring stroma across entire diagnostic biopsy WSIs (Mann Whitney U *p* = 0.043). Poor outcome patients had an event within 3 years, while Good outcome patients did not. **(E)** Patient pCR status has a much weaker association with Good and Poor RFS outcome.

Fig. 7A shows the association of spatial TILseg scoring with RFS using multivariate Cox regression, accounting for tumor stage at diagnosis, for 0, 797, and 1594 µm^2^ of epithelial clusters filtered out. These correspond to zero, three, and five connected kernels (50 × 50 pixels) of epithelium class predictions by our 3CC tissue classifier. Supp. Fig. 1 illustrates that for most patients, the optimal spatial subset of stroma is more enriched with sTILs than the cumulative stroma across the biopsy slide. As such, it is more predictive of patient prognosis to score the inflammation immediately around TNBC tumors (within 50 µm) than TILs far away. Supp. Figs. 2C and 2D show that by scoring a subset of the stroma, we are sampling a smaller area of stroma (TILseg score denominator) and sTILs (TILseg score numerator) as opposed to the whole slide, but the relative decrease in the stromal area is larger than that of the sTILs- resulting in a higher TILseg score (sTIL density) for most patients. Supp. Fig. 2A and 2B show that for a large spatial subset of stroma (within 2 µm of epithelial clusters with no size-based filtering), there is little difference in stroma or sTIL area compared to global scoring. Essentially, we are sampling almost the entire slide with this set of spatial parameters, compared to a much more exclusive sample of the slide with the optimal spatial parameters (within 50 µm of epithelial clusters, at least 797 µm²). Supp. Fig. 3 illustrates the effect of filtering out other sizes of epithelial clusters. This, combined with the optimal distance thresholding in the stroma as discussed above, yielded in the most prognostic sTILs isolation resulting in a strong association with patient RFS (HR = 0.65, 95% CI [0.44-0.96], *p* = 0.0289). This optimal spatial subset specifically captures lymphocytes in immediate proximity to substantial epithelial structures while excluding both the distant stromal lymphocytes and small epithelial fragments that may represent isolated epithelial cells or small ducts. Filtering out epithelial clusters less than 797 µm^2^ results in the optimal removal of noisy epithelial predictions for smoother TILseg scoring in spatial subsets of stroma (Fig. 7A).

The Kaplan Meier curve in Fig. 7B illustrates a more significant stratification of patient RFS (log-rank *p* = 0.00918) with spatial TILseg scoring (stroma within 50 µm of epithelial clusters at least 797 µm^2^) than with global TILseg scoring (log-rank *p* = 0.0191, Fig. 5D). This further substantiates the importance of scoring a subset of sTILs within stromal regions closer to epithelium in diagnostic biopsies for predicting patient risk. The cutoff thresholds for both global and spatial TILseg scoring were chosen for the optimal log-rank *p*-value. Given the established association between pCR and favorable outcomes (42), we examined whether pCR status could also stratify patients by RFS in our cohort. We found that even though patients with pCR showed a trend towards improved RFS, pCR status was not able to stratify patients based on RFS in a statistically significant manner (Fig. 7C, log-rank *p* = 0.106).

To directly compare the discriminatory power of spatial TILseg and global TILseg scoring, we stratified patients by good versus poor RFS outcomes and compared TILseg distributions (Fig. 7D). Spatial scoring resulted in a larger separation between the two groups, patients with good outcomes exhibiting higher sTIL scores than patients with poor outcomes (Mann-Whitney U p = 0.0062). On the other hand, global scoring showed weaker discrimination (Mann-Whitney U p = 0.043) with a higher overlap in the distributions. The improved effect size and statistical significance with spatial subset scoring reinforce that immune infiltration within 50 microns of epithelial clusters captures a more clinically relevant immune interaction than global immune assessment.

Again, pCR status could not stratify patients into good and poor RFS groups (Fig. 7E, Fisher’s Exact *p* = 0.14). Spatial TILseg scores, which were evaluated at the time of diagnosis (prior to any treatment), have a significantly greater potential for predicting recurrence risk response post-neoadjuvant therapy than pCR status, which was determined after the treatment. Although pCR was not significantly associated with recurrence risk in this study, it remains important to evaluate a diagnostic biomarker’s ability to predict pCR status before neoadjuvant therapy. (3,4,7,9–11) In the discovery cohort, nearly all (12/13) patients who achieved pCR experienced a recurrence-free survival longer than three years (Fig. 7E).

The computed TILseg scores were significantly associated with pCR rates after neoadjuvant chemotherapy in the discovery cohort (Fig. 8A). Spatial scoring of sTILs in the optimal stromal region (AUC = 0.72, Mann Whitney U *p* = 0.0065) is more significantly associated with pCR rates than scoring sTILs across the entire biopsy WSI (AUC = 0.67, Mann Whitney U *p* = 0.026), with both spatial and global scoring being statistically significant. To evaluate the generalizability of these findings, we applied identical spatial parameters to an independent validation cohort (n = 30, Fig. 8B). The associations for both spatial and global scoring with pCR trend towards statistical significance, with the spatial TILseg scores (AUC = 0.68, Mann Whitney U *p* = 0.053) being more significant than global TILseg scores (AUC = 0.65, Mann Whitney U *p* = 0.086). The box plots for spatial scoring reveal clear separation between groups, with pCR patients clustering in higher sTIL score ranges and non-pCR patients showing lower or more variable scores. Global scoring showed attenuated discrimination with greater overlap between pCR and non-pCR distributions. Importantly, the spatial scoring approach maintained clinically meaningful AUC values across both cohorts, suggesting potential utility as a pretreatment biomarker for identifying patients most likely to achieve pCR.

**Figure 8.**
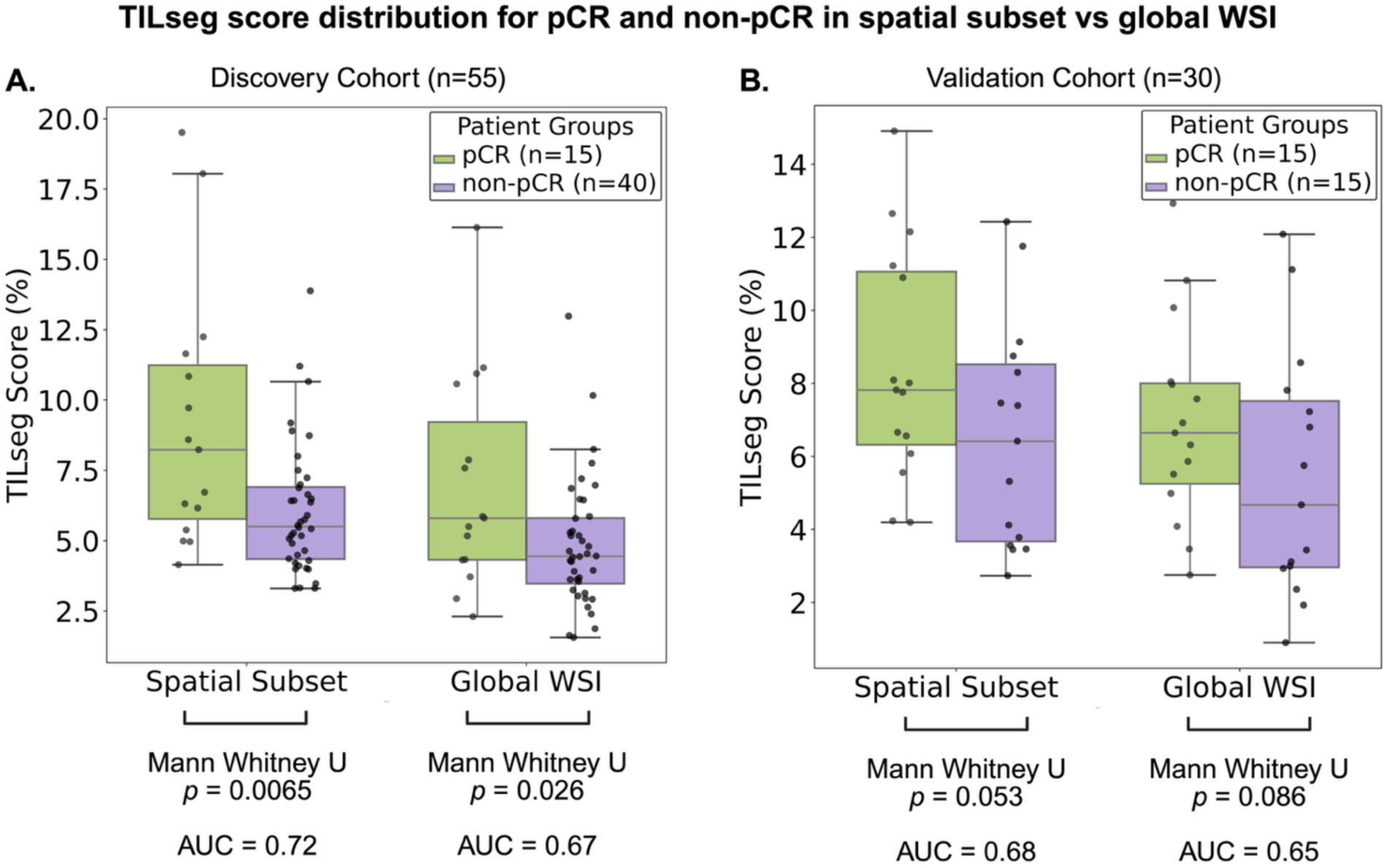
TILseg scoring of diagnostic biopsies as a predictive biomarker for pCR status. **(A)** TILseg scoring of the discovery cohort in a spatial subset of stroma within 50 µm of epithelial clusters that are at least 797 µm^2^ results in a significantly better stratified distribution of the scores (AUC=0.72, Mann Whitney U *p*=0.0065) across patient groups that achieved pCR and those that did not, compared to scoring stroma across entire diagnostic biopsy WSIs (AUC=0.67, Mann Whitney U *p*=0.026). **(B)** Similar to the discovery cohort, TILseg scoring of the independent validation cohort in the spatial subset (AUC=0.68, Mann Whitney U *p*=0.053) has a stronger association with pCR status than the global biopsy (AUC=0.65, Mann Whitney U *p*=0.086), with both trending towards statistical significance.

## DISCUSSION

The development and validation of TILseg, a computational pipeline that operates as an end-to-end framework for continuous sTIL scoring and sTIL spatial analysis in WSIs, represents a significant advance in computational pathology for sTILs profiling in breast cancer. TILseg combines automated segmentation of key histological features with principles from the International TILs Working Group to generate biologically relevant sTIL scores, expressed as the proportion of stromal area occupied by lymphocytic infiltration. Our results demonstrate that TILseg produces scores that achieve high concordance with expert pathologists, while also significantly enhancing the prognostic value of sTILs in predicting recurrence free survival and pCR status from diagnostic biopsies in TNBC patients. This establishes TILseg as a reliable surrogate for manual assessment while offering substantial advantages in objectivity, reproducibility, and scalability for clinical implementation and large-scale research applications. The elimination of subjective visual estimation and limited regional sampling inherent in manual scoring (43,44) addresses a persistent barrier to adoption of sTILs as a routine clinical biomarker, despite strong evidence supporting their prognostic and predictive value in breast cancer. Since, our pipeline samples the entire whole slide image for TILs scoring, it will enable clinicians to incorporate and visualize the spatial heterogeneity of sTILs across the entire WSI and utilize it in a quantitative and standardized manner to improve patient prognostication.

Another notable strength of our pipeline is its ability to adapt to a range of staining and imaging variations across multiple patient cohorts. With a few annotations from a handful of WSIs, our base model can be fine-tuned to generate predictions with a reasonably high level of accuracy in an independent dataset. This is supported by the high correlation coefficients between TILseg scoring and standardized manual scoring we observed in the independent validation cohort, implying there is a consistent and highly agreeable relationship between TILseg and clinician scoring. However, the computational outputs were notably lower with scores ranging from 0% up to 12% whereas pathologist scores ranged from 0% to 90%. This difference may be attributed to varied sampling where TILseg systematically accounts for the entire tissue area while pathologists may prioritize areas with higher levels of infiltration in their scores or differ in the way they calculate the area occupied by sTILs as a fraction of the total stromal compartment, introducing sampling bias which can lead to varying diagnoses. Beyond sampling, intrinsic differences between humans and automated algorithms in how they assess medical images may also contribute to this score range disparity. While TILseg analyzes these images at a pixel-level scale, pathologists will synthesize contextual, morphological, and spatial information holistically and subjectively. The granularity of a computational model likely inflates the denominator of the sTIL score as it comprehensively accounts for the stroma across the entire tissue area, thus resulting in deflated scores. This is further supported by the fact that even though a cutoff of 10% is most used for manual TILs scoring, a cutoff of 3.4% came out to be most prognostic for the computational scores produced by the TILseg pipeline, and a cutoff of 6.4% for the spatial TILseg pipeline.

Multivariate Cox regression demonstrates that TILseg-derived global sTIL scores from diagnostic core needle biopsies have strong, independent prognostic value for RFS in TNBC patients. Kaplan-Meier analysis revealed that TILseg significantly stratified patients into low and high-risk groups at optimal cutoffs where manual scoring could not achieve statistical significance, a finding further confirmed by Mann-Whitney U test. These results indicate that automated sTIL quantification captures clinically meaningful immune responses from pre-treatment biopsies that manual methods may overlook, highlighting the prognostic potential of computational immune profiling. In addition to global scoring, our spatial TILseg approach enables us to systematically look at how spatial subsets of lymphocytes affect survival outcomes. The systematic evaluation of spatial compartments revealed that sTILs quantified within 50 µm of substantial epithelial clusters (> 797 µm^2^) provided superior prognostic stratification (log-rank p = 0.00918). The predictive value of the spatial TILseg score for pathologic complete response to NAC, both in the discovery (AUC = 0.72) and independent validation (AUC = 0.68) cohorts represents a particularly clinically actionable finding. These findings align with emerging understanding that the spatial architecture of the tumor immune microenvironment carries biological and clinical information beyond simple cell counts. (16–19) The biologically- motivated spatial filtering implemented in TILseg captures a functionally relevant immunological niche that conventional global scoring methods cannot isolate. This work also suggests that spatial TILs scoring in diagnostic biopsies could inform treatment decision making providing complementary prognostic information.

The consistent performance across cohorts supports the generalizability of the TILseg scoring, though validation in larger, prospectively collected cohorts will be essential before clinical implementation. Since TILs are not unique to TNBC, TILseg could be adapted to other clinical breast cancer subtypes and immune active cancers, such as colorectal cancer, non-small cell lung cancer, and melanoma (45–48), where systematic quantification and assessment of their spatial heterogeneity has the potential to guide therapeutic decision-making. Additionally, while our pipeline accurately identifies TILs based on morphological criteria, it does not distinguish between functionally distinct lymphocyte populations such as cytotoxic CD8+ T cells, Tregs, or B cells that may have different prognostic implications. Integration with multiplexed immunofluorescence platforms or spatial transcriptomics could address this limitation, enabling spatially resolved phenotypic characterization. In conclusion, TILseg establishes a robust foundation for spatially-informed immune profiling in breast cancer, demonstrating that computational approaches can not only replicate expert pathologist performance but also extract novel prognostic and predictive features capturing clinically relevant spatial organization of the tumor-immune-microenvironment.

## Data Availability

All code produced is available online at https://github.com/Shachi-Mittal-Lab/TILseg.

## ACKNOWLEDGEMENTS & FUNDING

We thank the National Institutes of Health for funding through K99CA293004, P30CA015704, R01CA248192. We acknowledge funding from ASCO/CCF Hayden Family Foundation Young Investigator Award in Breast Cancer, University of Washington Institute of Medical Data Science Pilot Award, University of Washington Chemical Engineering Startup Grant, and the Roger E. Moe Fellowship in Multidisciplinary Breast Cancer Care. This research was also supported by the Biostatistics Shared Resource of the Fred Hutch/University of Washington/Seattle Children’s Cancer Consortium (P30CA015704).

## CODE AVAILABILITY

The guidance and code for global and spatial sTIL scoring is available in the GitHub repository at https://github.com/Shachi-Mittal-Lab/TILseg. All code is available under a non-commercial license.

## AUTHOR’S CONTRIBUTIONS

S.M, L.C, and A.S conceptualized and designed the research. A.K, S.P, M.K, L.S, D.R, S.D, J.S, and L.K provided the pathology data and annotations. L.C, A.S, K.H, and M.R carried out the development of the computational pipeline for sTIL scoring. L.C, A.S, and D.H carried out the statistical and survival analysis. S.M, L.C and A.S wrote the manuscript. A.K, S.P, L.S, M.K, and D.H provided additional clinical insights and reviewed/edited the manuscript.

