## Supplemental Data for "TILseg: Automated Whole Slide-Level Spatial Scoring of Tumor-Infiltrating Lymphocytes Reveals Prognostic Patterns in Triple Negative Breast Cancer"

SUPPLEMENTARY

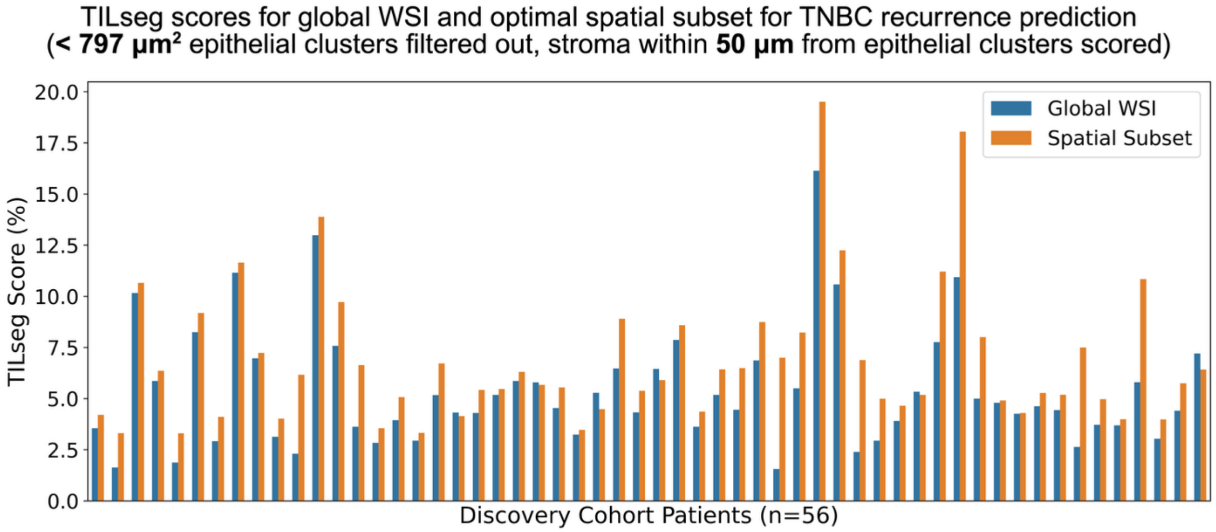


**Figure S1.** Global TILseg scores for the discovery cohort patients. The density of sTILs in stroma (TILseg score) is higher for most patients in a spatial subset within 50 μm of epithelial clusters that are at least 797 $\mu m^2$ than across the entire biopsy WSI.

**A.** TILseg score **denominator** for global WSI and a **large** spatial subset of stroma (no size-based epithelia noise filtering, stroma within **2 mm** from epithelial clusters scored)

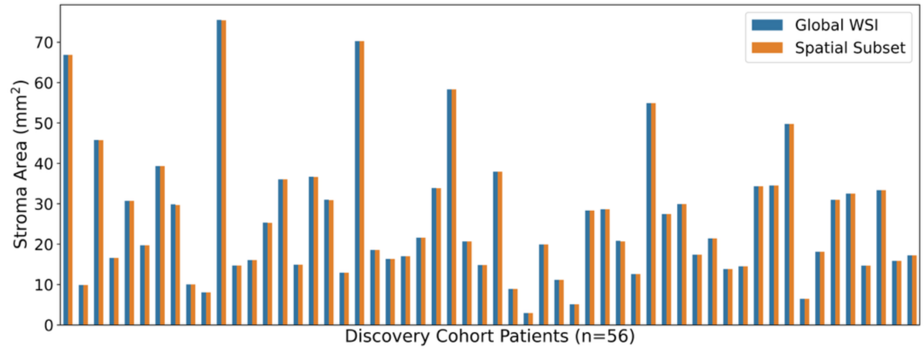


**B.** TILseg score **numerator** for global WSI and a **large** spatial subset of stroma (no size-based epithelia noise filtering, stroma within **2 mm** from epithelial clusters scored)

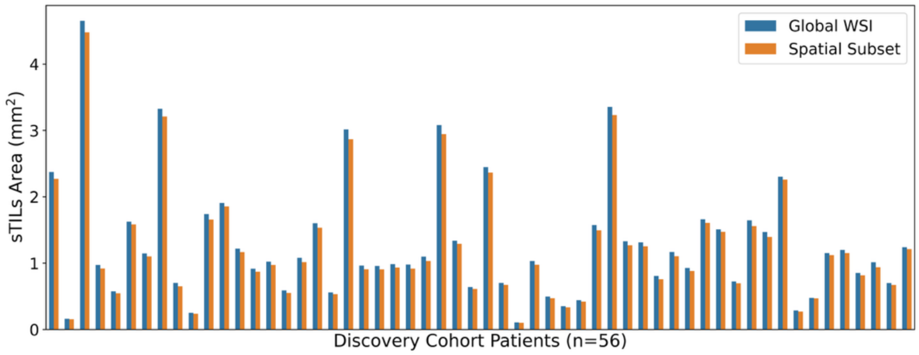


**C.** TILseg score **denominator** for global WSI and a **small** spatial subset of stroma (**< 797 μm²** epithelial clusters filtered out, stroma within **50 μm** from epithelial clusters scored)

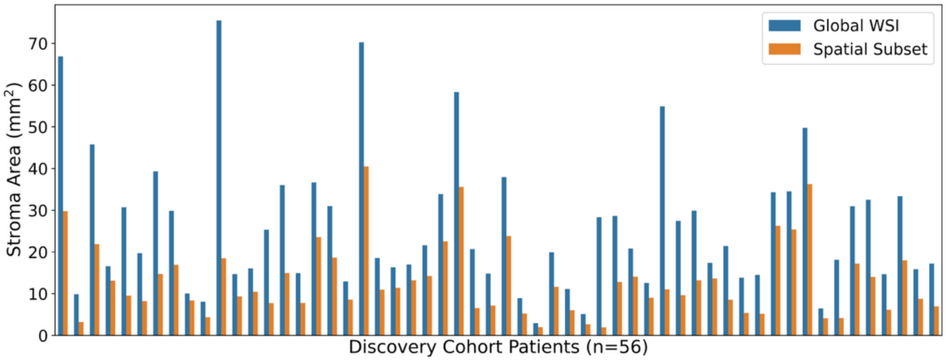


**D.** TILseg score **numerator** for global WSI and a **small** spatial subset of stroma (**< 797 μm²** epithelial clusters filtered out, stroma within **50 μm** from epithelial clusters scored)

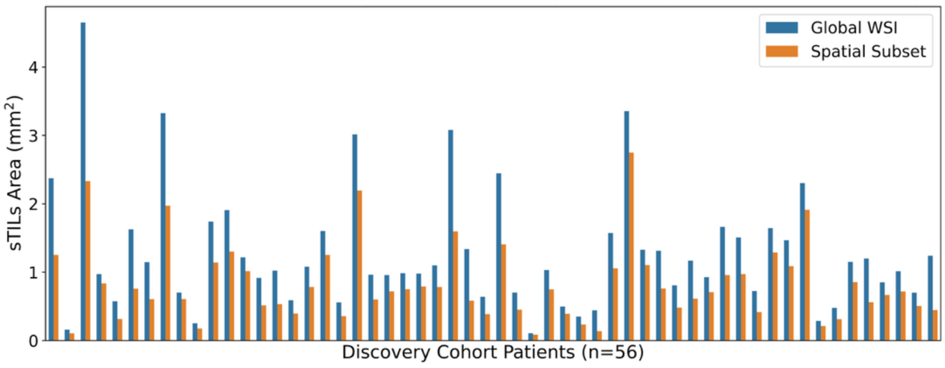

**Figure S2.** sTILs and stroma area scored in global and spatial TILseg scoring for the discovery cohort patients. **(A)** The stromal area (TILseg score denominator) is almost the same for all patients in a large spatial subset within 2 mm of epithelia with no size-based filtering of epithelial noise than across the entire biopsy WSI. **(B)** The sTILs area (TILseg score numerator) is almost the same for all patients in a large spatial subset within 2 mm of epithelia with no size-based filtering of epithelial noise than across the entire biopsy WSI. **(A)** and **(B)** show that scoring the large spatial subset is almost equivalent to scoring the entire WSI. (**C**) The stroma area scored (TILseg score denominator) is much lower for most patients in a spatial subset within 50 µm of epithelial clusters that are at least 797 µm$^2$ than across the entire biopsy WSI. **(D)** sTILs area (TILseg score numerator) is much lower for most patients in a spatial subset within 50 µm of epithelial clusters that are at least 797 µm$^2$ than across the entire biopsy WSI.

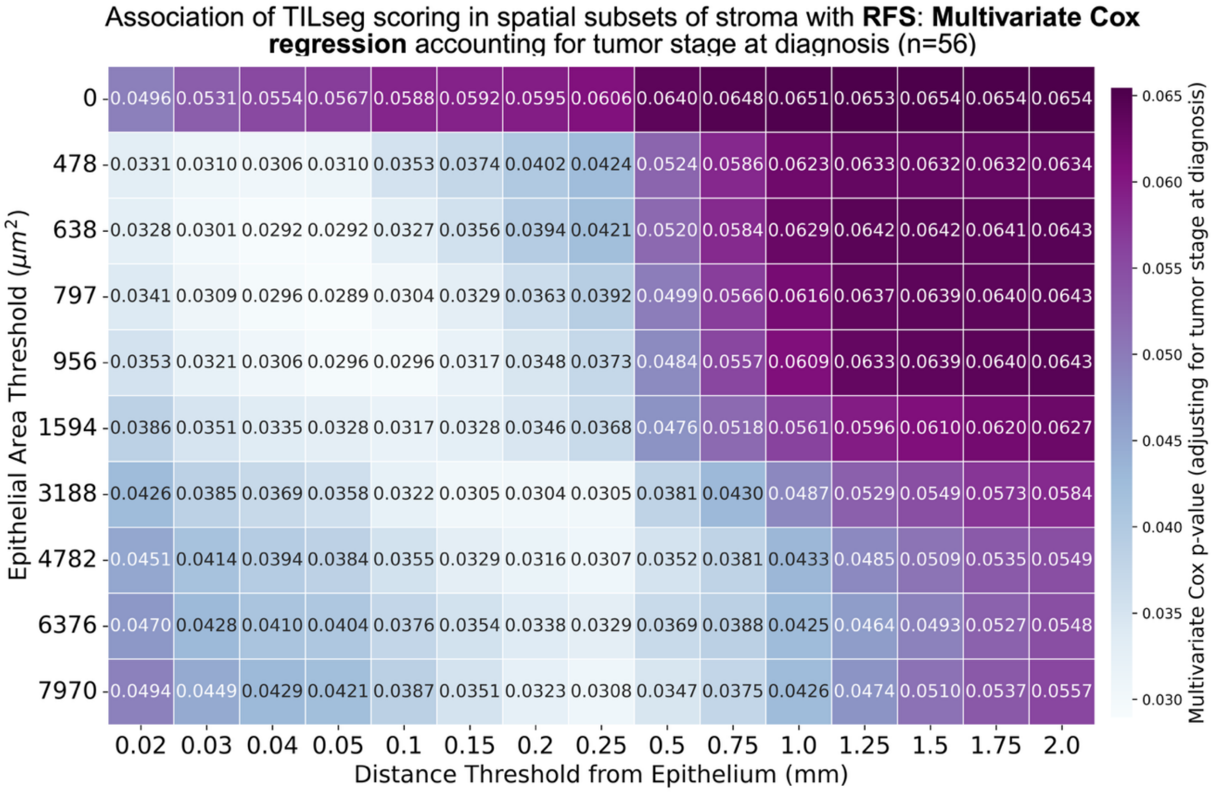


**Figure S3**. Multivariate Cox proportional hazards regression p values for TILseg scoring in the spatial subsets of stroma around epithelia in the diagnostic biopsies of the Discovery Cohort (n = 56 TNBC patients). The heatmap shows the iterations of TILseg scoring in various spatial stroma subsets. The columns denote the distance from epithelial clusters within which stroma was scored for sTILs and the rows indicate the size below which epithelial clusters were considered as noise and removed from the scoring analysis. On the y-axis, 0, 478, 638, 797, 956, 1594, 3188, 4782, 6376, and 7970 µm$^2$ correspond to 0, 3, 4, 5, 6, 10, 20, 30, and 50 kernels (50 × 50 pixel) of 3CC predictions, respectively. A lower p value in the heatmap illustrates a more significant association of the spatial subset TILseg score with patient RFS independently of the tumor stage at diagnosis.

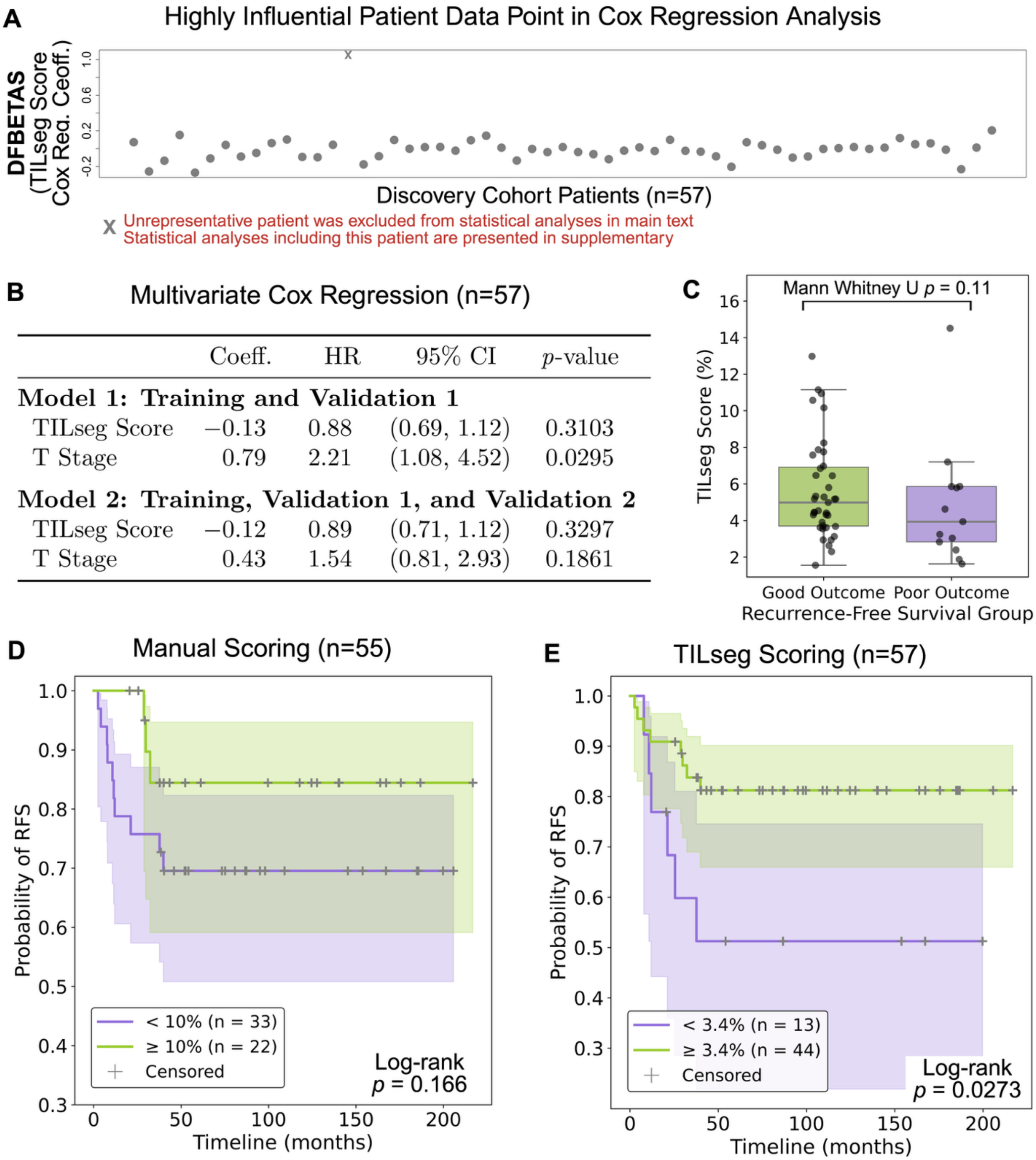


| | Coeff. | HR | 95% CI | $p$-value |
|---|---|---|---|---|
| **Model 1: Training and Validation 1** | | | | |
| TILseg Score | −0.13 | 0.88 | (0.69, 1.12) | 0.3103 |
| T Stage | 0.79 | 2.21 | (1.08, 4.52) | 0.0295 |
| **Model 2: Training, Validation 1, and Validation 2** | | | | |
| TILseg Score | −0.12 | 0.89 | (0.71, 1.12) | 0.3297 |
| T Stage | 0.43 | 1.54 | (0.81, 2.93) | 0.1861 |

**Figure S4.** Analyses including the highly influential patient. (**A**) A multivariate Cox regression model was fitted for the entire Discovery Cohort (n=57), accounting for tumor stage at diagnosis. One patient was identified as unrepresentative and highly influential in the regression analysis using DFBETAS. This patient was excluded from the analyses in the main text, and is included in the analyses shown in Figure S4 and S5. (**B**) Multivariate Cox regression results for the training and validation I subsets (n = 49) after adjusting for clinical tumor stage. A second multivariate Cox regression model was fitted on WSIs in the training, validation I, and validation II samples (n = 57). (**C**) Patients in the discovery cohort who had recurred within 3 years (poor outcome, n = 13) had significantly lower median TILseg scores than patients who had not recurred within 3 years (good outcome, n = 39). Kaplan-Meier stratification of RFS is shown for the discovery cohort by (**D**) manual scoring (n = 55) and (**E**) TILseg scores (n = 57).

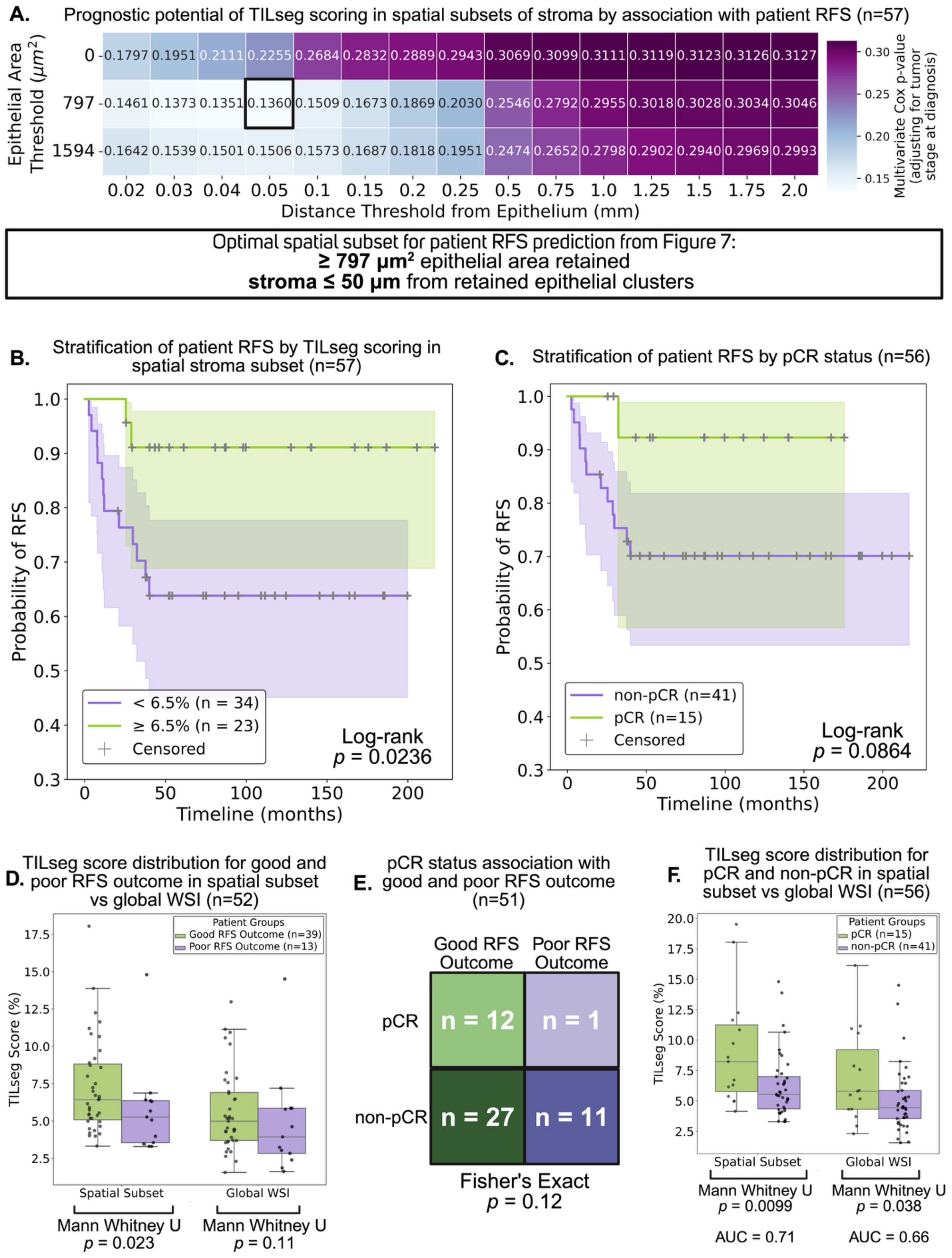
A.
Prognostic potential of TILseg scoring in spatial subsets of stroma by association with patient RFS (n=57)
Epithelial Area Threshold ($\mu m^2$)
0 - 0.1797 0.1951 0.2111 0.2255 0.2684 0.2832 0.2889 0.2943 0.3069 0.3099 0.3111 0.3119 0.3123 0.3126 0.3127
797 - 0.1461 0.1373 0.1351 0.1360 0.1509 0.1673 0.1869 0.2030 0.2546 0.2792 0.2955 0.3018 0.3028 0.3034 0.3046
1594 - 0.1642 0.1539 0.1501 0.1506 0.1573 0.1687 0.1818 0.1951 0.2474 0.2652 0.2798 0.2902 0.2940 0.2969 0.2993
0.02 0.03 0.04 0.05 0.1 0.15 0.2 0.25 0.5 0.75 1.0 1.25 1.5 1.75 2.0
Distance Threshold from Epithelium (mm)
Multivariate Cox p-value (adjusting for tumor stage at diagnosis)
0.30 0.25 0.20 0.15
Optimal spatial subset for patient RFS prediction from Figure 7:
≥ 797 µm² epithelial area retained
stroma ≤ 50 µm from retained epithelial clusters
B.
Stratification of patient RFS by TILseg scoring in spatial stroma subset (n=57)
Probability of RFS
Timeline (months)
< 6.5% (n = 34)
≥ 6.5% (n = 23)
Censored
Log-rank
p = 0.0236
C.
Stratification of patient RFS by pCR status (n=56)
Probability of RFS
Timeline (months)
non-pCR (n=41)
pCR (n=15)
Censored
Log-rank
p = 0.0864
D.
TILseg score distribution for good and poor RFS outcome in spatial subset vs global WSI (n=52)
TILseg Score (%)
Patient Groups
Good RFS Outcome (n=39)
Poor RFS Outcome (n=13)
Spatial Subset
Global WSI
Mann Whitney U
p = 0.023
Mann Whitney U
p = 0.11
E.
pCR status association with good and poor RFS outcome (n=51)
Good RFS Outcome
Poor RFS Outcome
pCR
n = 12
n = 1
non-pCR
n = 27
n = 11
Fisher's Exact
p = 0.12
F.
TILseg score distribution for pCR and non-pCR in spatial subset vs global WSI (n=56)
TILseg Score (%)
Patient Groups
pCR (n=15)
non-pCR (n=41)
Spatial Subset
Global WSI
Mann Whitney U
p = 0.0099
Mann Whitney U
p = 0.038
AUC = 0.71
AUC = 0.66

**Figure S5.** Analyses including the highly influential patient. **(A)** Multivariate Cox proportional hazards regression *p* values for spatial TILseg scoring in different subsets of stroma around epithelia using diagnostics biopsies from the Discovery Cohort (n=57) accounting for tumor stage at diagnosis. **(B)** Kaplan-Meier curve shows significant improvement in the stratification of patient RFS by spatial TILseg scoring using the optimal spatial stromal pocket and epithelial cluster size (log-rank test). **(C)** Patient pCR status has a weaker stratification potential of RFS. **(D)** TILseg scoring in the optimal spatial subset results in a significantly better stratified distribution of the scores across Good and Poor RFS patient groups than scoring stroma across entire diagnostic biopsy WSIs. Poor outcome patients had a recurrence within 3 years, while Good outcome patients did not. **(E)** Patient pCR status has a much weaker association with Good and Poor RFS outcomes. **(F)** TILseg scoring in the spatial subset results in a significantly better stratified distribution of the scores across patient groups that achieved pCR and those that did not, compared to scoring stroma across entire diagnostic biopsy WSIs.
